# Rapid loss of doxycycline effectiveness against gonorrhea after implementation of post-exposure prophylaxis in southern California: an observational study

**DOI:** 10.64898/2026.01.07.26343623

**Authors:** Matan Yechezkel, David Helekal, Banshri Kapadia, Vennis Hong, Magdalena E. Pomichowski, Iris Anne C. Reyes, Gregg S. Davis, Nicola F. Müller, Yonatan H. Grad, Sara Y. Tartof, Joseph A. Lewnard

**Author notes:** These authors contributed equally (Sara Y. Tartof, Joseph A. Lewnard). Corresponding authors: Sara Tartof, 100 S. Los Robles, Pasadena, California 91101, United States, Joseph Lewnard, 2121 Berkeley Way, Berkeley, California 94720, United States.

## Abstract

**Background:** Post-exposure prophylaxis with doxycycline (doxyPEP) within 72 hours after sex is recommended to prevent chlamydia, gonorrhea, and syphilis among United States men who have sex with men (MSM) and transgender women (TGW). However, concerns surround potential expansion of *Neisseria gonorrhoeae* lineages harboring antimicrobial resistance (AMR) in settings that implement doxyPEP.

**Methods:** We assessed the effectiveness of doxyPEP against chlamydia, gonorrhea, and syphilis among individuals receiving care from the Kaiser Permanente Southern California integrated healthcare system between 1 January, 2023 and 30 June, 2025. Primary analyses estimated treatment effectiveness by comparing history of doxyPEP prescription fills ≤90 days before positive or negative tests for infection with *Chlamydia trachomatis, N. gonorrhoeae*, and *Treponema pallidum*. We evaluated changes in treatment effectiveness against gonorrhea over time and in association with the proportion of local *N. gonorrhoeae* isolates harboring *tetM*, a plasmid-borne gene conferring high-level tetracycline resistance.

**Results:** Among 26,582 individuals meeting eligibility criteria, 2,262 (8·5%) received ≥1 doxyPEP fill during the study period. DoxyPEP fills occurred ≤90 days before 39 of 4,659 (0·8%) positive and 3,288 of 130,138 (2·5%) negative *C. trachomatis* tests; 212 of 6,539 (3·2%) positive and 3,112 of 128,238 (2·4%) negative *N. gonorrhoeae* tests; and 11 of 2,169 (0·5%) positive and 794 of 26,913 (3·0%) negative *T. pallidum* tests. Throughout the study period, treatment effectiveness of doxyPEP against chlamydia, gonorrhea, and syphilis was 66·5% (53·6–75·9%), –1·8% (–18·5 to 12·5%), and 60·7% (28·3–78·5%), respectively. Treatment effectiveness against gonorrhea declined from 42·3% (2·7–65·8%) before statewide doxyPEP implementation to –15·0% (–51·1 to 11·4%) from January–June, 2025, in association with rapid expansion of *tetM* prevalence among circulating *N. gonorrhoeae* lineages. Treatment effectiveness during periods with *tetM* detected among 20-29·9% of *N. gonorrhoeae* isolates was 47·2% (4·1–70·3%), declining to −8·5% (−33·2 to 11·2%) during periods with *tetM* detected among ≥50% of isolates.

**Conclusions:** We report rapid loss of doxyPEP effectiveness against gonorrhea within the first year after doxyPEP implementation among MSM and TGW in southern California, associated with expansion of high-level tetracycline resistance in *N. gonorrhoeae*. This outcome, alongside persisting effectiveness against chlamydia and syphilis, may inform risk-benefit considerations for doxyPEP implementation strategies.

**Funding:** US Centers for Disease Control and Prevention

## INTRODUCTION

In June, 2024, the United States Centers for Disease Control & Prevention (CDC) recommended that healthcare providers counsel men who have sex with men (MSM) and transgender women (TGW) about doxycycline post-exposure prophylaxis (doxyPEP), and consider prescribing doxyPEP to these patients as a strategy to prevent chlamydia, gonorrhea, and syphilis.^1^ This recommendation encompasses patients diagnosed with STIs in the preceding 12 months, as well as patients who did not receive STI diagnoses but will participate in sexual activities known to increase their likelihood of STI exposure. Individuals receiving doxyPEP are recommended to take 200mg doxycycline orally as soon as possible within 72 hours after condomless anal, vaginal, or oral sex to prevent infections. Prior to US-wide recommendations, doxyPEP was recommended locally in San Francisco beginning in October, 2022,^2^ and statewide throughout California in April, 2023.^3^

Pre-implementation randomized controlled trials demonstrated that doxyPEP reduced incidence of chlamydia and syphilis among MSM and TGW by 70-89% and 73-87%, respectively.^4–6^ Whereas doxyPEP also reduced gonorrhea incidence by 55% among US trial participants,^6^ protection was not evident in a trial undertaken in France, in which 78% of incident, culture-positive *N. gonorrhoeae* infections showed reduced susceptibility to tetracycline.^5^ Secondary analyses of trial data from France reported elevated prevalence of *tetM*—a plasmid-borne gene associated with high-level tetracycline resistance^7^—in *N. gonorrhoeae* isolates obtained from doxyPEP recipients in comparison to nonrecipients.^8^ Genetic elements associated with reduced susceptibility to cephalosporins, penicillins, and quinolones often co-occur with *tetM*, and were likewise detected more commonly in isolates from doxyPEP recipients than non-recipients in the trial. In the United States, the proportion of *N. gonorrhoeae* isolates carrying *tetM* increased from <10% in 2020 to >30% by the first quarter of 2024,^9^ underscoring concerns that doxyPEP implementation could contribute to AMR selection in *N. gonorrhoeae*.^10^ Among MSM receiving doxyPEP between November, 2022 and December, 2023 in northern California, incidence of chlamydia and syphilis declined by 79-80%, whereas incidence of gonorrhea declined only 12%.^11^

To better understand the effectiveness of doxyPEP against each of these STIs, we conducted a retrospective study of effectiveness among individuals receiving healthcare from Kaiser Permanente Southern California (KPSC), a large, integrated healthcare system serving 4·8 million members (~21% of the regional population). We evaluated differences in the protective effectiveness of doxyPEP against gonorrhea in association with calendar time and changes in the prevalence of *tetM* among local *N. gonorrhoeae* isolates.

## METHODS

### Setting

The KPSC healthcare system is an integrated, membership-based pre-paid, closed-network organization providing comprehensive care to members across virtual, outpatient, emergency department, and inpatient settings. Electronic healthcare records (EHRs) link all aspects of in-network care including diagnoses, prescriptions, laboratory testing, and clinical notes via unique patient identifiers, while out-of-network care is captured through insurance claim reimbursements; collectively, this enables near-complete ascertainment of members’ healthcare receipt. Members are enrolled in KPSC health plans via employer-sponsored, self-paid, and publicly-subsidized insurance schemes, and broadly reflect the regional population in terms of racial/ethnic diversity, socioeconomic characteristics, and prevalence of common chronic conditions.^12^

### Study population

Our study population comprised all KPSC members assigned male sex at birth, born on or before 30 September, 2008, and who were living with HIV infection or receiving HIV pre- or post-exposure prophylaxis (PrEP/PEP) at any time between 1 July, 2021 and 30 June, 2025 (**Table S1**; **Table S2**). Study follow-up commenced from the latest of the following dates: 1 January, 2023, one year after the date of health plan entry, or members’ 16^th^ birthday. Follow-up continued through the earliest of: 30 June, 2025, death, or disenrollment (allowing for lapses in membership lasting up to 45 days).

### Outcomes

We defined chlamydia and gonorrhea infections as the first positive detection of *C. trachomatis* and *N. gonorrhoeae* by nucleic acid amplification testing of specimens from any anatomical site in a 30-day period; we excluded data from repeat positive tests within 30 days, when tests may have been undertaken on a confirmatory basis or as tests of cure for previously-treated infections. We defined incident syphilis diagnoses as a new reactive plasma reagin test result or 4-fold increase in rapid plasma reagin titer relative to readings obtained ≥365 days prior, and excluded positive syphilis test results received <365 after prior incident infections.^13^

### Statistical analysis

Primary analyses used a test-negative design to assess treatment effectiveness of doxyPEP against laboratory-confirmed chlamydia, gonorrhea, and syphilis within ≤90 days, 91-180 days, and >180 days after a prescription fill among all cohort members. This framework was expected to mitigate confounding that could arise from differences in STI testing frequency among doxyPEP recipients and non-recipients.^14^ We defined doxyPEP fills as prescription dispenses including a ≥30-day supply of doxycycline, dispensed as either single 200mg tablets or 2×100mg tablets. We used logistic regression models to compute adjusted odds ratios (aORs) comparing history of doxyPEP fills ≤90, 91-180, or >180 days before testing, versus no history of doxyPEP fills within the study period, among individuals testing positive or negative for each infection. We defined treatment effectiveness as (1 − aOR) × 100%. Models adjusted for confounding variables specified *a priori* including individuals’ age group; race/ethnicity; cisgender or transgender identity (ascertained via natural language processing from structured and unstructured EHR fields^15^); HIV infection status or receipt of HIV PrEP/PEP in the prior 180 days; prior-year history of STI testing and STI diagnoses; history of JYNNEOS vaccination; commercial or non-commercial source of insurance coverage; community-level socioeconomic status, as measured by neighborhood deprivation index;^16^ other antibiotic use in the preceding 30 days; and calendar month of testing (categorizations of continuous covariates are presented in **Table 1**). Models included cluster-robust standard errors to account for repeated sampling of individuals across multiple tests, and rolled data across multiple tests within the same month into a single observation. We repeated analyses in subgroups defined based on STI risk (STI diagnoses received in the preceding 6 or 12 months) and based on testing frequency (receiving ≥2 or ≥4 tests in the preceding 12 months, per CDC guidelines for MSM at high risk^17^).

**Table 1:** Characteristics of individuals testing positive and those testing negative for each infection.

| Characteristics |  | Chlamydia |  | Gonorrhea |  | Syphilis |  |
| --- | --- | --- | --- | --- | --- | --- | --- |
|  |  | Negative, n (%) | Positive, n (%) | Negative, n (%) | Negative, n (%) | Negative, n (%) | Positive, n (%) |
|  |  | N=130,138 | N=4,659 | N=128,238 | N=6,539 | N=26,913 | N=2,169 |
| Age group<sup>1</sup> | 16-25 years | 15,876 (12.2%) | 859 (18.4%) | 15,471 (12.1%) | 1,256 (19.2%) | 2,002 (7.4%) | 255 (11.8%) |
|  | 26-35 years | 43,407 (33.4%) | 1,871 (40.2%) | 42,501 (33.1%) | 2,770 (42.4%) | 8,201 (30.5%) | 671 (30.9%) |
|  | 36-45 years | 29,407 (22.6%) | 1,023 (22.0%) | 28,912 (22.5%) | 1,513 (23.1%) | 6,420 (23.9%) | 500 (23.1%) |
|  | ≥46 years | 41,448 (31.8%) | 906 (19.4%) | 41,354 (32.2%) | 1,000 (15.3%) | 10,290 (38.2%) | 743 (34.3%) |
|  | Median (interquartile range) | 37 (30-50) | 33 (28-42) | 37 (30-51) | 33 (27-40) | 40 (32-53) | 38 (30-52) |
| Race/ethnicity | White, non-Hispanic | 43,140 (33.1%) | 1,123 (24.1%) | 42,555 (33.2%) | 1,703 (26.0%) | 8,027 (29.8%) | 456 (21.0%) |
|  | Black, non-Hispanic | 10,777 (8.3%) | 431 (9.3%) | 10,554 (8.2%) | 653 (10.0%) | 2,514 (9.3%) | 265 (12.2%) |
|  | Hispanic, any race | 54,347 (41.8%) | 2,285 (49.0%) | 53,545 (41.8%) | 3,078 (47.1%) | 12,685 (47.1%) | 1,149 (53.0%) |
|  | Asian/Pacific Islander, non-Hispanic | 11,799 (9.1%) | 440 (9.4%) | 11,708 (9.1%) | 527 (8.1%) | 1,916 (7.1%) | 153 (7.1%) |
|  | Other/Unknown | 10,075 (7.7%) | 380 (8.2%) | 9,876 (7.7%) | 578 (8.8%) | 1,771 (6.6%) | 146 (6.7%) |
| Gender identity | No evidence of transgender identity | 128,192 (98.5%) | 4,581 (98.3%) | 126,293 (98.5%) | 6,460 (98.8%) | 26,565 (98.7%) | 2,133 (98.3%) |
|  | Transgender identity recorded | 1,946 (1.5%) | 78 (1.7%) | 1,945 (1.5%) | 79 (1.2%) | 348 (1.3%) | 36 (1.7%) |
| Health insurance source | Commercial plan | 111,869 (86.0%) | 4,086 (87.7%) | 110,155 (85.9%) | 5,783 (88.4%) | 22,516 (83.7%) | 1,741 (80.3%) |
|  | Others/Unknown | 18,269 (14.0%) | 573 (12.3%) | 18,083 (14.1%) | 756 (11.6%) | 4,397 (16.3%) | 428 (19.7%) |
| Neighborhood deprivation index | Quartile 1 (least deprived) | 32,709 (25.1%) | 1,021 (21.9%) | 32,229 (25.1%) | 1,500 (22.9%) | 6,800 (25.3%) | 473 (21.8%) |
|  | Quartile 2 | 32,577 (25.0%) | 1,109 (23.8%) | 32,161 (25.1%) | 1,521 (23.3%) | 6,774 (25.2%) | 506 (23.3%) |
|  | Quartile 3 | 32,493 (25.0%) | 1,204 (25.8%) | 32,047 (25.0%) | 1,645 (25.2%) | 6,727 (25.0%) | 555 (25.6%) |
|  | Quartile 4 (most deprived) | 32,359 (24.9%) | 1,325 (28.4%) | 31,801 (24.8%) | 1,873 (28.6%) | 6,612 (24.6%) | 635 (29.3%) |
| HIV infection status or prevention | HIV infection | 15,185 (11.7%) | 477 (10.2%) | 15,089 (11.8%) | 572 (8.7%) | 4,386 (16.3%) | 539 (24.9%) |
|  | Pre/Post-exposure prophylaxis within 180 days | 76,725 (59.0%) | 2,600 (55.8%) | 75,296 (58.7%) | 4,022 (61.5%) | 14,054 (52.2%) | 447 (20.6%) |
| Mpox vaccination status | 0 JYNNEOS doses | 60,737 (46.7%) | 2,189 (47.0%) | 59,904 (46.7%) | 3,016 (46.1%) | 11,827 (43.9%) | 1,336 (61.6%) |
|  | ≥1 JYNNEOS dose | 69,401 (53.3%) | 2,470 (53.0%) | 68,334 (53.3%) | 3,523 (53.9%) | 15,086 (56.1%) | 833 (38.4%) |
| Prior-year gonorrhea tests | 0-2 tests | 35,933 (27.6%) | 1,154 (24.8%) | 35,840 (27.9%) | 1,229 (18.8%) | 7,584 (28.2%) | 1,448 (66.8%) |
|  | 3-5 tests | 65,278 (50.2%) | 2,120 (45.5%) | 64,358 (50.2%) | 3,046 (46.6%) | 12,437 (46.2%) | 551 (25.4%) |
|  | 6-11 tests | 27,072 (20.8%) | 1,283 (27.5%) | 26,271 (20.5%) | 2,076 (31.7%) | 6,270 (23.3%) | 160 (7.4%) |
|  | ≥12 tests | 1,855 (1.4%) | 102 (2.2%) | 1,769 (1.4%) | 188 (2.9%) | 622 (2.3%) | 10 (0.5%) |
| Prior chlamydia diagnoses | 0 diagnoses | 102,783 (79.0%) | 2,906 (62.4%) | 101,741 (79.3%) | 3,920 (59.9%) | 19,439 (72.2%) | 1,809 (83.4%) |
|  | 1 diagnosis | 19,601 (15.1%) | 1,127 (24.2%) | 19,087 (14.9%) | 1,650 (25.2%) | 4,926 (18.3%) | 276 (12.7%) |
|  | ≥2 diagnoses | 7,754 (6.0%) | 626 (13.4%) | 7,410 (5.8%) | 969 (14.8%) | 2,548 (9.5%) | 84 (3.9%) |
| Prior gonorrhea diagnoses | 0 diagnoses | 99,625 (76.6%) | 2,836 (60.9%) | 98,923 (77.1%) | 3,551 (54.3%) | 18,630 (69.2%) | 1,790 (82.5%) |
|  | 1 diagnosis | 20,549 (15.8%) | 1,156 (24.8%) | 20,006 (15.6%) | 1,683 (25.7%) | 5,214 (19.4%) | 277 (12.8%) |
|  | ≥2 diagnoses | 9,964 (7.7%) | 667 (14.3%) | 9,309 (7.3%) | 1,305 (20.0%) | 3,069 (11.4%) | 102 (4.7%) |
| Prior syphilis diagnosis | No | 105,164 (80.8%) | 3,173 (68.1%) | 103,892 (81.0%) | 4,427 (67.7%) | 16,508 (61.3%) | 1,231 (56.8%) |
|  | Yes | 24,974 (19.2%) | 1,486 (31.9%) | 24,346 (19.0%) | 2,112 (32.3%) | 10,405 (38.7%) | 938 (43.2%) |
| Recent (30d) antibiotic treatment | No treatment | 115,350 (88.6%) | 4,069 (87.3%) | 114,224 (89.1%) | 5,145 (78.7%) | 23,632 (87.8%) | 1,900 (87.6%) |
|  | STI-related treatment | 2,864 (2.2%) | 414 (8.9%) | 2,521 (2.0%) | 768 (11.7%) | 439 (1.6%) | 139 (6.4%) |
|  | Treatment unrelated to STI | 11,924 (9.2%) | 176 (3.8%) | 11,493 (9.0%) | 626 (9.6%) | 2,842 (10.6%) | 130 (6.0%) |
| Other risk factors |  |  |  |  |  |  |  |
|  | Alcohol or drug abuse diagnosis | 15,407 (11.8%) | 598 (12.8%) | 15,066 (11.7%) | 937 (14.3%) | 3,835 (14.2%) | 407 (18.8%) |

Data summarize 26,582 unique individuals (26,377 tested for chlamydia, 26,376 tested for gonorrhea, and 7,213 tested for syphilis) among 33,118 meeting eligibility for study inclusion.

To explore potential consequences of resistance emergence, we also evaluated effect modification in the relationship between doxyPEP receipt and *N. gonorrhoeae* infection. We estimated treatment effectiveness over distinct periods, which we defined as: January–April, 2023 (prior to recommendations for statewide doxyPEP implementation^3^); May– December, 2023 (early implementation phase); and throughout each subsequent year included in follow-up (January– December, 2024 and January–June, 2025). We also examined effectiveness in relation to the proportion of *N. gonorrhoeae* isolates harboring *tetM* collected from patients presenting to sentinel sexual health clinics in three counties within the study region (Los Angeles, Orange, and San Diego). Primary analyses defined prevalence of *tetM* among circulating *N. gonorrhoeae* isolates collected from the Gonococcal Isolate Surveillance Project (GISP), which are obtained from males with new-onset urethral gonorrhea, using whole-genome sequence data, as described previously^9^ (**Text S1**). We repeated analyses using a broader dataset including isolates from GISP and from two other CDC-led projects: enhanced GISP, which collected *N. gonorrhoeae* from non-urethral sites, and Strengthening the United States Response to Resistant Gonorrhea, which focused on antibiotic-resistant *N. gonorrhoeae*. We estimated treatment effectiveness separately for periods when *tetM* was present in 20–29·9%, 30–39·9%, 40–49·9%, and ≥50% of local sequenced isolates. We also fit models defining time and *tetM* prevalence as continuous effect modifiers, comparing penalized fits across candidate models via the Bayesian Information Criterion; we used the resulting models to estimate time to loss of doxyPEP effectiveness against gonorrhea, defined as the first date at which aOR = 1 for the association of a positive gonorrhea test result with doxyPEP receipt in the preceding 90 days (**Text S2**).

As a secondary analysis, we evaluated changes in incidence rates of chlamydia, gonorrhea, and syphilis among the study cohort throughout follow-up from January 1, 2023 through June 30, 2025. We measured weekly incidence rates via Poisson regression models, defining counts of incident laboratory-confirmed infections as the outcome and the log-transformed eligible population size as an offset term. Models defined calendar week as a continuous term and included sine- and cosine transformations of calendar week, defined at a 1-year period length, to adjust for seasonality.

### Ethics

The Kaiser Permanente Southern California Institutional Review Board approved the study and provided a waiver of the requirement for informed consent for retrospective analyses of EHR data.

## RESULTS

The cohort meeting eligibility criteria comprised 33,118 individuals, among whom 26,582 received ≥1 STI test between 1 January, 2023 and 30 June, 2025 and were thus included in primary analyses (**Table S3**; **Figure 1**). Of these individuals, 2,262 (8·5%) received ≥1 doxyPEP fill. Median age at index among doxyPEP recipients was younger (34 years; interquartile range: 29–43) than among non-recipients (38 years; 30–53), and recipients were more likely than non-recipients to be Hispanic (1,102 [48·7%] of 2,262 versus 9,932 [40·8%] of 24,320), more likely to have commercial insurance coverage (1,982 [87·6%] versus 20,463 [84·1%]), more likely to have received ≥1 JYNNEOS (mpox) vaccine dose (1,578 [69·8%] versus 12,461 [51·2%]), and less likely to be HIV positive (157 [6·9%] versus 4,005 [16·5%]). Recipients of doxyPEP were also more likely than non-recipients to have received chlamydia (303 [13·6%] versus 2,068 [8·5%]), gonorrhea (356 [15·7%] versus 2,223 [9·1%]), or syphilis (416 [18·4%] versus 3,765 [15·5%]) diagnoses in the year before cohort entry.

**Figure 1:**
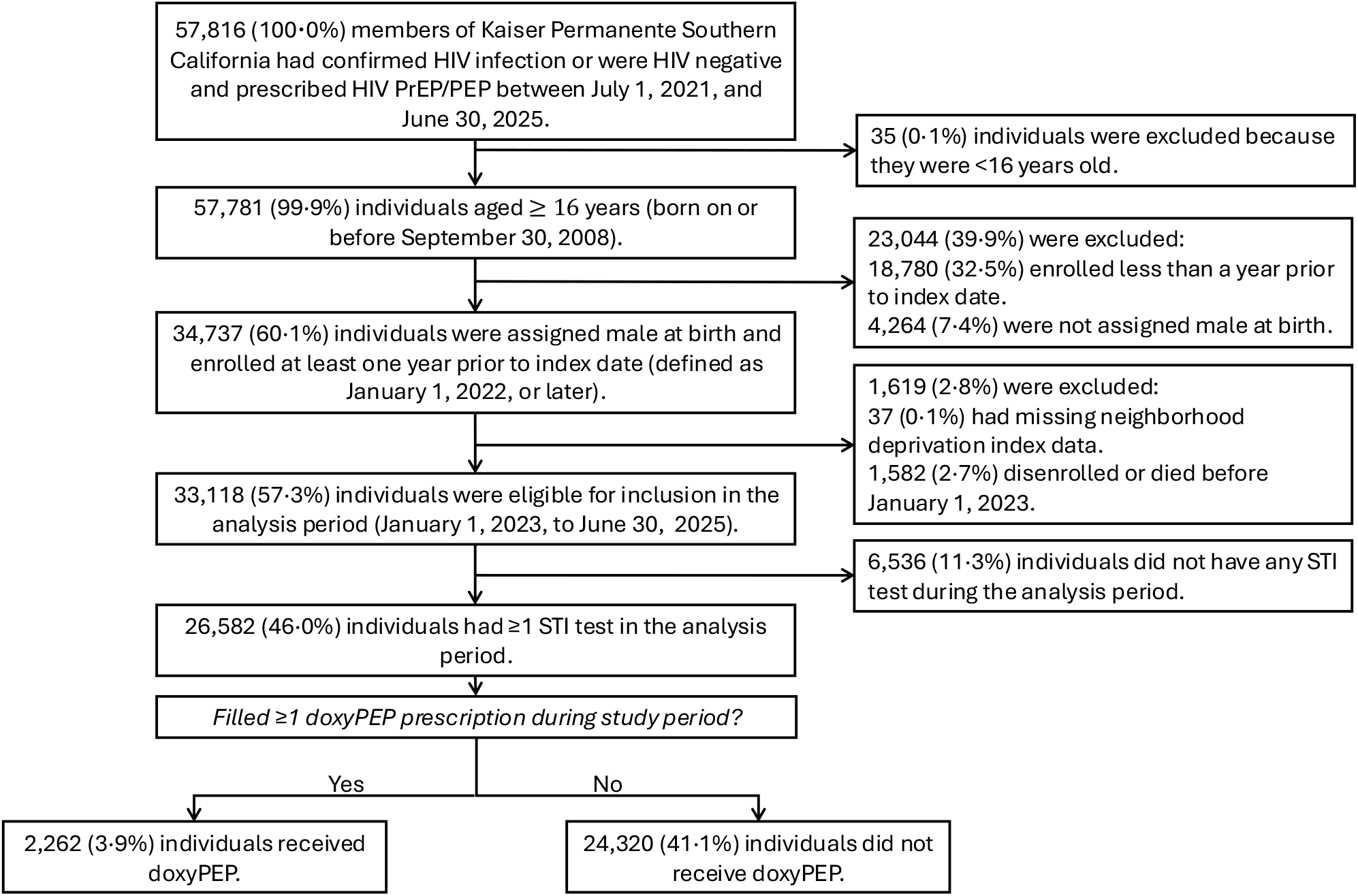
Study flowchart. We detail the numbers of individuals meeting eligibility criteria and included in the study as doxyPEP recipients or non-recipients.

Follow-up captured 134,797 eligible chlamydia tests among 26,377 unique individuals, 134,777 eligible gonorrhea tests among 26,376 unique individuals, and 29,082 eligible syphilis tests among 7,213 unique individuals (**Table 1**). In total, 4,659 (3·5%) chlamydia, 6,539 (4·9%) gonorrhea, and 2,169 (7·5%) syphilis tests yielded positive results. Individuals testing positive for each infection were generally younger than those testing negative, more likely to be Hispanic, more likely to live in neighborhoods experiencing socioeconomic deprivation, and received greater numbers of STI tests and STI diagnoses in the preceding year.

Overall, 8,877 (6·8%) of 130,138 negative chlamydia tests and 175 (3·8%) of 4,659 positive chlamydia tests were preceded by any doxyPEP fill; 3,288 (2·5%) negative tests and 39 (0·8%) positive tests were preceded by a doxyPEP fill within 90 days before testing (**Table 2**). Treatment effectiveness of doxyPEP for prevention of chlamydia was 66·5% (53·6–75·9%) within 90 days after prescription fill, 53·4% (33·4–67·3%) within 91-180 days, and 16·2% (–4·7 to 32·9%) within >180 days. Across all syphilis tests, 2,089 (7·8%) of 26,913 negative tests and 51 (2·4%) of 2,169 positive tests were preceded by any doxyPEP fill, with 794 (3·0%) negative tests and 11 (0·5%) positive tests preceded by a doxyPEP fill within 90 days before testing. DoxyPEP treatment effectiveness against syphilis was 60·7% (28·3–78·5%) within 90 days after prescription fill, 41·8% (–5·9 to 68·0%) within 91-180 days, and 41·2% (10·9–61·2%) within >180 days. We obtained similar estimates of treatment effectiveness against chlamydia and syphilis in analyses restricted to individuals with recent STI diagnoses and higher frequency of STI testing within the preceding 6 or 12 months (**Table S4, Table S5**).

**Table 2:**
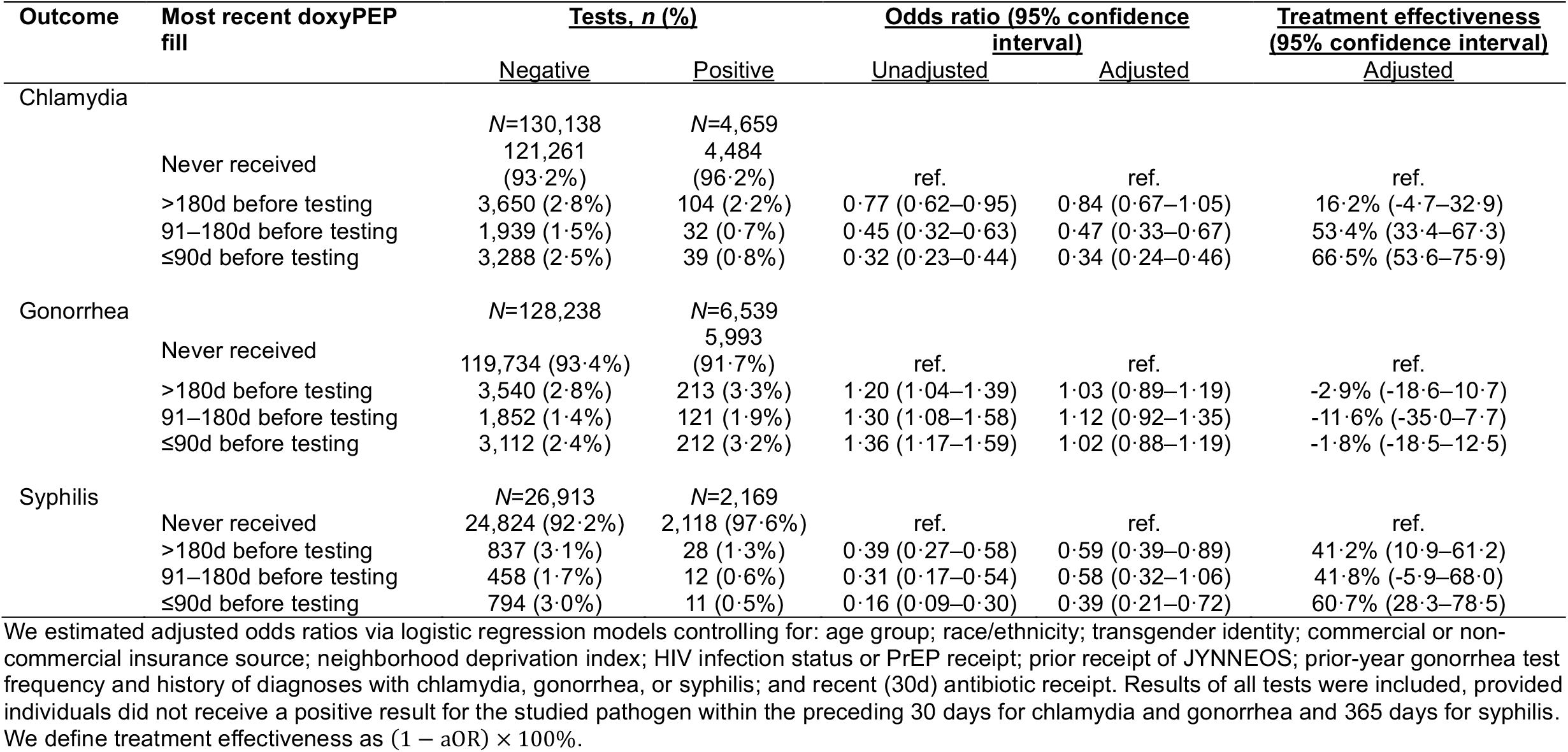
Effectiveness of doxyPEP against chlamydia, gonorrhea, and syphilis.

| Outcome | Most recent doxyPEP fill | Tests, n (%) |  | Odds ratio (95% confidence interval) |  | Treatment effectiveness (95% confidence interval) |
| --- | --- | --- | --- | --- | --- | --- |
|  |  | Negative | Positive | Unadjusted | Adjusted | Adjusted |
| Chlamydia |  |  |  |  |  |  |
|  |  | N=130,138 | N=4,659 |  |  |  |
|  | Never received | 121,261 (93.2%) | 4,484 (96.2%) | ref. | ref. | ref. |
|  | >180d before testing | 3,650 (2.8%) | 104 (2.2%) | 0.77 (0.62–0.95) | 0.84 (0.67–1.05) | 16.2% (-4.7–32.9) |
|  | 91–180d before testing | 1,939 (1.5%) | 32 (0.7%) | 0.45 (0.32–0.63) | 0.47 (0.33–0.67) | 53.4% (33.4–67.3) |
|  | ≤90d before testing | 3,288 (2.5%) | 39 (0.8%) | 0.32 (0.23–0.44) | 0.34 (0.24–0.46) | 66.5% (53.6–75.9) |
| Gonorrhea |  |  |  |  |  |  |
|  |  | N=128,238 | N=6,539 |  |  |  |
|  | Never received | 119,734 (93.4%) | 5,993 (91.7%) | ref. | ref. | ref. |
|  | >180d before testing | 3,540 (2.8%) | 213 (3.3%) | 1.20 (1.04–1.39) | 1.03 (0.89–1.19) | -2.9% (-18.6–10.7) |
|  | 91–180d before testing | 1,852 (1.4%) | 121 (1.9%) | 1.30 (1.08–1.58) | 1.12 (0.92–1.35) | -11.6% (-35.0–7.7) |
|  | ≤90d before testing | 3,112 (2.4%) | 212 (3.2%) | 1.36 (1.17–1.59) | 1.02 (0.88–1.19) | -1.8% (-18.5–12.5) |
| Syphilis |  |  |  |  |  |  |
|  |  | N=26,913 | N=2,169 |  |  |  |
|  | Never received | 24,824 (92.2%) | 2,118 (97.6%) | ref. | ref. | ref. |
|  | >180d before testing | 837 (3.1%) | 28 (1.3%) | 0.39 (0.27–0.58) | 0.59 (0.39–0.89) | 41.2% (10.9–61.2) |
|  | 91–180d before testing | 458 (1.7%) | 12 (0.6%) | 0.31 (0.17–0.54) | 0.58 (0.32–1.06) | 41.8% (-5.9–68.0) |
|  | ≤90d before testing | 794 (3.0%) | 11 (0.5%) | 0.16 (0.09–0.30) | 0.39 (0.21–0.72) | 60.7% (28.3–78.5) |

We estimated adjusted odds ratios via logistic regression models controlling for: age group; race/ethnicity; transgender identity; commercial or non-commercial insurance source; neighborhood deprivation index; HIV infection status or PrEP receipt; prior receipt of JYNNEOS; prior-year gonorrhea test frequency and history of diagnoses with chlamydia, gonorrhea, or syphilis; and recent (30d) antibiotic receipt. Results of all tests were included, provided individuals did not receive a positive result for the studied pathogen within the preceding 30 days for chlamydia and gonorrhea and 365 days for syphilis. We define treatment effectiveness as $(1 - aOR) \times 100\%$ .

Overall, 8,504 (6·6%) of 128,238 negative gonorrhea tests and 546 (8·3%) of 6,539 positive gonorrhea tests were preceded by any doxyPEP fill, with 3,112 (2·4%) negative tests and 212 (3·2%) positive tests occurring within ≤90 days after a fill (**Table 2**). Effectiveness of doxyPEP against gonorrhea was –1·8% (–18·5 to 12·5%), –11·6% (–35·0 to 7·7%), and –2·9% (–18·6 to 10·7%) within ≤90, 91-180, and >180 days after individuals’ most recent prescription fill, and declined over time in association with rapid increases in *tetM* prevalence following statewide doxyPEP implementation (**Figure 2**). Among *N. gonorrhoeae* isolates from GISP in the study region, *tetM* was detected in 21·7% (15/69) of those collected from January to April, 2023, before doxyPEP implementation; this proportion rose to 40·5% (60/148) of specimens collected from May to December, 2023, and 59·1% (75/127) of specimens collected from January to December, 2024 (**Table S6**). Similar trends were observed in combined GISP and non-GISP isolates (**Table S6; Figure S1**). Effectiveness of doxyPEP against gonorrhea within ≤90 days after a prescription fill was 42·3% (2·7–65·8%) from January to April, 2023; following statewide doxyPEP implementation, effectiveness declined to 28·0% (–1·1 to 48·6%) from May to December, 2023, –4·0% (–25·3 to 13·7%) throughout 2024, and –15·0% (–51·1 to 11·4%) in January–June, 2025 (**Table 3**). Similarly, effectiveness declined with increasing *tetM* prevalence among circulating *N. gonorrhoeae*. Whereas effectiveness of doxyPEP against gonorrhea within ≤90 days after a prescription fill was 47·2% (4·1–70·3%) during months with 20-20·9% *tetM* prevalence, this estimate declined to 35·5% (1·1–57·2%), 21·1% (−3·7 to 39·4%), and −8·5% (−33·2 to 11·2%) during months with 30·0–39·9%, 40·0–49·9%, and ≥50.0% *tetM* prevalence, respectively. We estimated that doxyPEP ceased to be effective in preventing gonorrhea by 2 June, 2024 (interquartile range: 1 April to 31 July, 2024) and by 18 March, 2024 (interquartile range: 28 December, 2023 to 14 September, 2024), respectively, in analyses modeling time and *tetM* prevalence as modifiers of treatment effectiveness (**Figure 3**). Results were consistent in analyses including non-GISP *N. gonorrhoeae* isolates for *tetM* prevalence estimation (**Figure S2; Table S7**)

**Table 3:** Effectiveness of doxyPEP against gonorrhea, by time and *tetM* prevalence (derived from GISP *N. gonorrhoeae* isolates).

| Prevalence range of *tetM* or period | Most recent doxyPEP fill | Tests, n (%) |  | Odds ratio (95% confidence interval) |  | Treatment effectiveness (95% confidence interval) |
| --- | --- | --- | --- | --- | --- | --- |
|  |  | Negative | Positive | Unadjusted | Adjusted | Adjusted |
| *tetM* prevalence 20–29.9% |  | *N*=16,178 | *N*=807 |  |  |  |
|  | Never received | 15,923 (98.4%) | 798 (98.9%) | ref. | ref. | ref. |
|  | >180d before testing | 119 (0.7%) | 4 (0.5%) | 0.88 (0.53–1.45) | 0.77 (0.45–1.31) | 23.3% (–31.4–55.0) |
|  | 91–180d before testing | 55 (0.3%) | 3 (0.4%) | 1.45 (0.78–2.73) | 1.26 (0.65–2.43) | –26.3% (–143.1–34.6) |
|  | ≤90d before testing | 81 (0.5%) | 2 (0.2%) | 0.84 (0.49–1.43) | 0.53 (0.30–0.96) | 47.2% (4.1–70.3) |
| *tetM* prevalence 30–39.9% |  | *N*=17,504 | *N*=993 |  |  |  |
|  | Never received | 17,113 (97.8%) | 975 (98.2%) | ref. | ref. | ref. |
|  | >180d before testing | 187 (1.1%) | 9 (0.9%) | 0.96 (0.67–1.37) | 0.82 (0.56–1.20) | 18.4% (–20.1–44.5) |
|  | 91–180d before testing | 66 (0.4%) | 3 (0.3%) | 1.46 (0.93–2.29) | 1.24 (0.77–1.99) | –24.0% (–98.6–22.9) |
|  | ≤90d before testing | 138 (0.8%) | 6 (0.6%) | 0.99 (0.68–1.45) | 0.65 (0.43–0.99) | 35.5% (1.1–57.2) |
| *tetM* prevalence 40–49.9% |  | *N*=20,995 | *N*=1,186 |  |  |  |
|  | Never received | 20,188 (96.2%) | 1,125 (94.9%) | ref. | ref. | ref. |
|  | >180d before testing | 300 (1.4%) | 21 (1.8%) | 1.05 (0.82–1.33) | 0.87 (0.67–1.13) | 13.2% (–12.6–32.8) |
|  | 91–180d before testing | 149 (0.7%) | 12 (1.0%) | 1.46 (1.09–1.97) | 1.22 (0.89–1.66) | –21.8% (–66.0–10.6) |
|  | ≤90d before testing | 358 (1.7%) | 28 (2.4%) | 1.17 (0.91–1.50) | 0.79 (0.61–1.04) | 21.1% (–3.7–39.4) |
| *tetM* prevalence ≥50% |  | *N*=47,409 | *N*=2,377 |  |  |  |
|  | Never received | 43,556 (91.9%) | 2,111 (88.8%) | ref. | ref. | ref. |
|  | >180d before testing | 1,471 (3.1%) | 87 (3.7%) | 1.21 (0.98–1.49) | 0.96 (0.78–1.19) | 4.5% (–18.7–22.5) |
|  | 91–180d before testing | 909 (1.9%) | 70 (2.9%) | 1.47 (1.16–1.87) | 1.18 (0.94–1.52) | –18.4% (–52.5–6.5) |
|  | ≤90d before testing | 1,473 (3.1%) | 109 (4.6%) | 1.52 (1.26–1.84) | 1.09 (0.89–1.33) | –8.5% (–33.2–11.2) |
| Pre-implementation period: January–April, 2023 |  | *N*=16,178 | *N*=807 |  |  |  |
|  | Never received | 15,923 (98.4%) | 798 (98.9%) | ref. | ref. | ref. |
|  | >180d before testing | 119 (0.7%) | 4 (0.5%) | 0.90 (0.56–1.44) | 0.80 (0.49–1.31) | 19.8% (–30.9–51.0) |
|  | 91–180d before testing | 55 (0.3%) | 3 (0.4%) | 1.49 (0.83–2.71) | 1.33 (0.72–2.43) | –32.7% (–143.4–27.8) |
|  | ≤90d before testing | 81 (0.5%) | 2 (0.2%) | 0.91 (0.57–1.48) | 0.58 (0.34–0.97) | 42.3% (2.7–65.8) |
| Early implementation phase: May–December, 2023 |  | *N*=33,863 | *N*=1,920 |  |  |  |
|  | Never received | 32,898 (97.1%) | 1,861 (96.9%) | ref. | ref. | ref. |
|  | >180d before testing | 416 (1.2%) | 24 (1.3%) | 1.00 (0.73–1.37) | 0.85 (0.62–1.18) | 14.8% (–17.9–38.4) |
|  | 91–180d before testing | 157 (0.5%) | 8 (0.4%) | 1.49 (1.02–2.20) | 1.28 (0.86–1.91) | –27.8% (–90.6–14.2) |
|  | ≤90d before testing | 392 (1.2%) | 27 (1.4%) | 1.09 (0.80–1.50) | 0.72 (0.51–1.01) | 28.0% (–1.1–48.6) |
| Mature implementation phase: January–December, 2024 |  | *N*=51,045 | *N*=2,636 |  |  |  |
|  | Never received | 47,959 (94.0%) | 2,350 (89.1%) | ref. | ref. | ref. |
|  | >180d before testing | 1,542 (3.0%) | 93 (3.5%) | 1.19 (0.98–1.44) | 0.94 (0.78–1.16) | 5.7% (–15.6–22.4) |
|  | 91–180d before testing | 967 (1.9%) | 77 (2.9%) | 1.47 (1.18–1.86) | 1.20 (0.95–1.52) | –20.1% (–51.8–4.8) |
|  | ≤90d before testing | 1,577 (3.1%) | 116 (4.4%) | 1.47 (1.23–1.76) | 1.04 (0.86–1.25) | –4.0% (–25.3–13.7) |
| Mature implementation phase: January–June, 2025 |  | *N*=26,152 | *N*=1,176 |  |  |  |
|  | Never received | 22,954 (87.8%) | 984 (83.7%) | ref. | ref. | ref. |
|  | >180d before testing | 1463 (5.6%) | 92 (7.8%) | 1.49 (1.20–1.86) | 1.23 (0.99–1.55) | –23.1% (–54.6–1.2) |
|  | 91–180d before testing | 673 (2.6%) | 33 (2.8%) | 1.17 (0.83–1.69) | 0.96 (0.67–1.40) | 4.1% (–40.2–33.1) |
|  | ≤90d before testing | 1062 (4.0%) | 67 (5.7%) | 1.48 (1.16–1.92) | 1.15 (0.89–1.51) | –15.0% (–51.1–11.4) |

We estimated adjusted odds ratios via logistic regression models controlling for: age group; race/ethnicity; transgender identity; commercial or non-commercial insurance source; neighborhood deprivation index; HIV infection status or PrEP receipt; prior receipt of JYNNEOS; prior-year gonorrhea test frequency and history of diagnoses with chlamydia, gonorrhea, or syphilis; and recent (30d) antibiotic receipt. Results of all tests were included, provided individuals did not receive a positive result for the studied pathogen within the preceding 30 days. We define treatment effectiveness as $(1 - aOR) \times 100\%$ . Analyses allowing effect modification by *tetM* prevalence included 24,542 unique individuals, due to data availability extending only through December, 2024.

**Figure 2:**
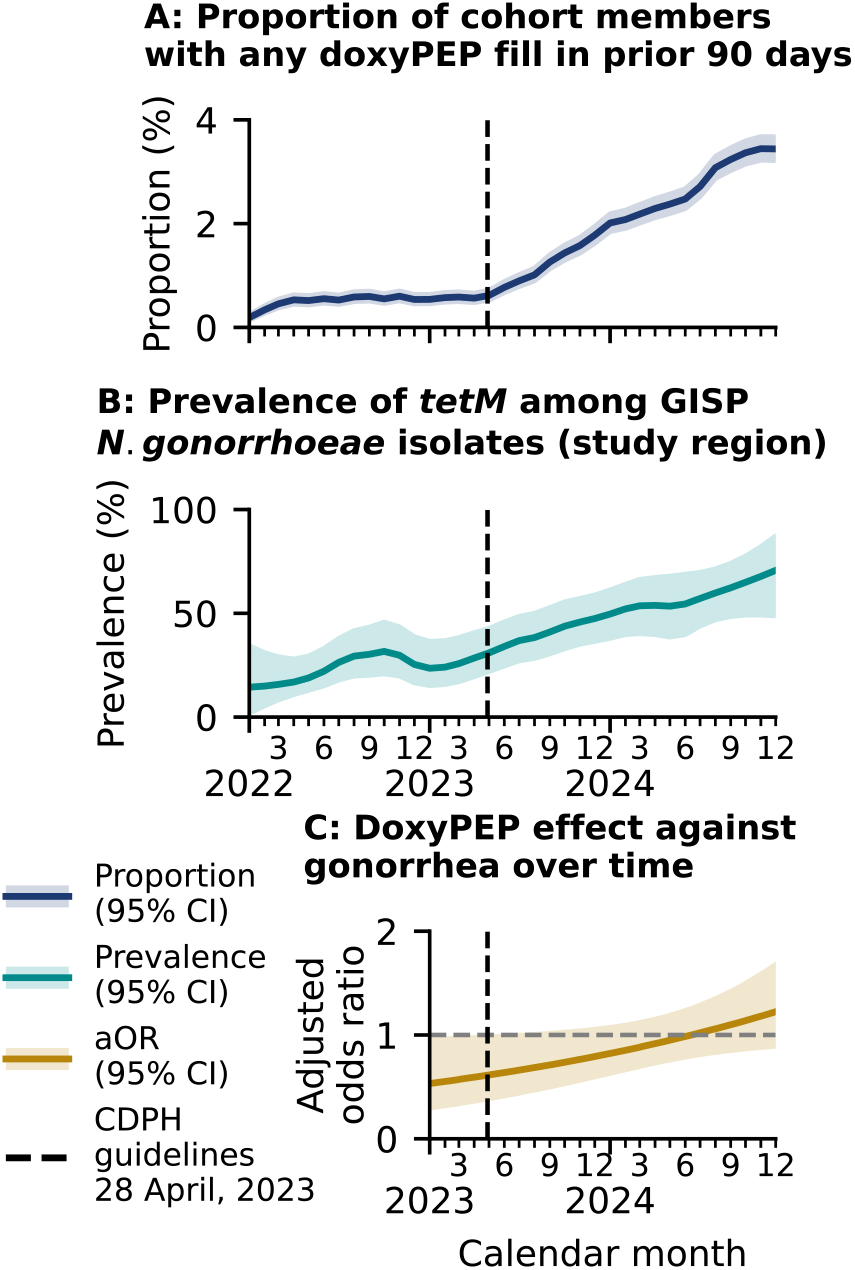
DoxyPEP uptake, *tetM* expansion, and time-varying effectiveness of doxyPEP against gonorrhea. We illustrate (**a**) the proportion of cohort members with a history of filling any doxyPEP prescription within the preceding 90 days, from January, 2022 through December, 2024; (**b**) the proportion of GISP *N. gonorrhoeae* isolates within the study region with *tetM* detected, shown with weighted LOESS smoothing (**Table S6**); and (**c**) the adjusted odds ratio of a doxyPEP prescription fill in the preceding 90 days among individuals testing positive or negative for *N. gonorrhoeae* infection, defining time as a continuous interaction term based on comparisons of fit for alternative models (**Table S7**). Shaded areas delineate 95% confidence intervals around point estimates (lines). We present numerical estimates of doxyPEP effectiveness during time intervals before and after doxyPEP implementation, by year, in **Table 3**.

**Figure 3:**
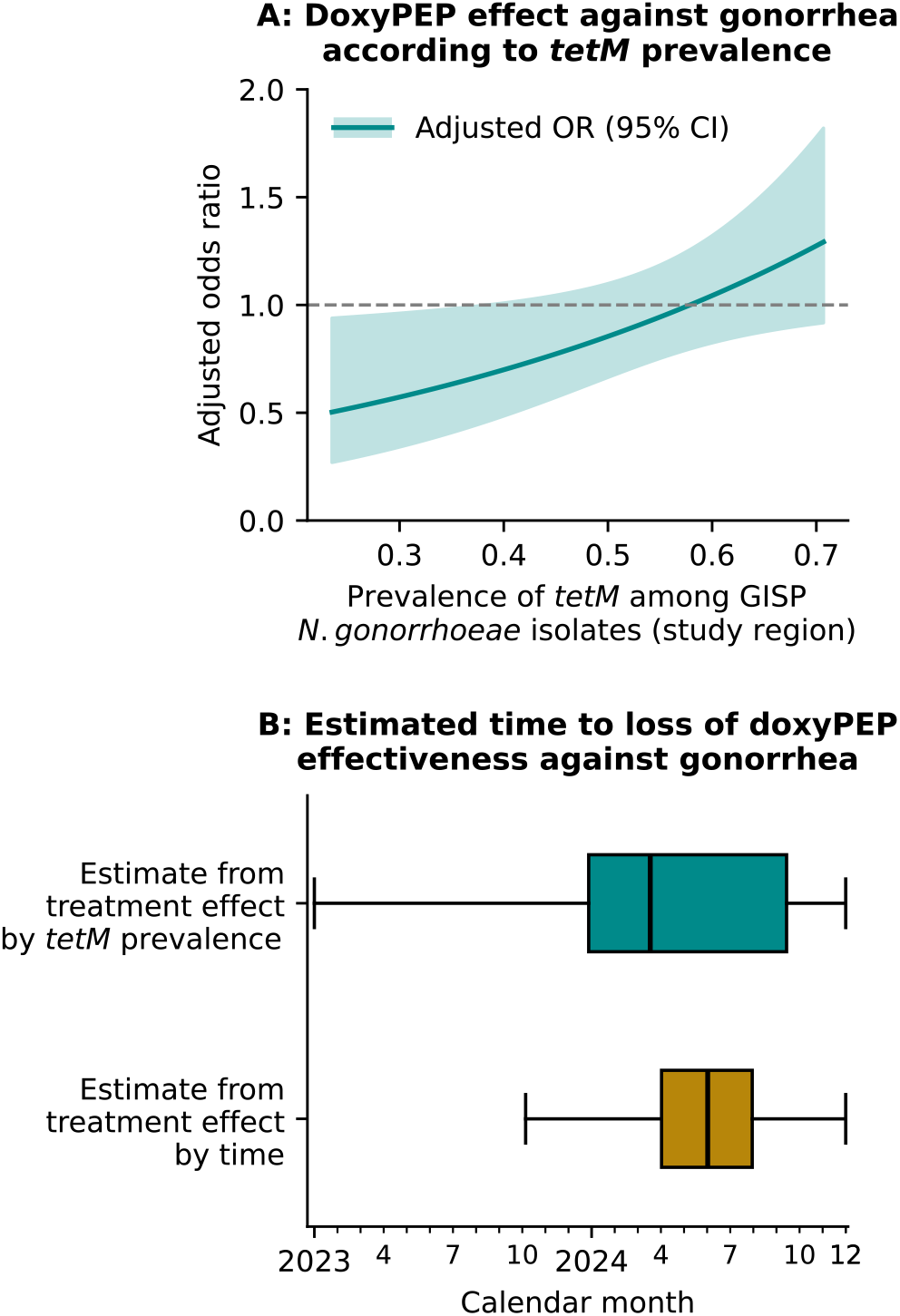
Variation in doxyPEP effectiveness with *tetM* prevalence, and time to loss of effect. We illustrate (**a**) the adjusted odds ratio of a doxyPEP prescription fill in the preceding 90 days among individuals testing positive or negative for *N. gonorrhoeae* infection, defining monthly *tetM* prevalence among GISP *N. gonorrhoeae* isolates within the study region as a continuous, linear interaction term based on comparisons of fit for alternative models (**Table S8**). Shaded areas delineate 95% confidence intervals around point estimates. Below, we illustrate (**b**) estimates of the time of loss of doxyPEP effectiveness against gonorrhea, defined as the last date *t* after with aOR(*t*)<1. We project estimates from models defining aOR as a function of *tetM* prevalence (as plotted in panel **a**) and time (as plotted in **Figure 2c**). Shaded areas delineate interquartile ranges around median estimates (lines); outer lines extend to 95% confidence intervals.

Between January, 2023 and June, 2025, annualized incidence of laboratory-confirmed chlamydia within the study population declined from 12·8 to 4·6 cases per 100 person-years at risk (**Figure 4**), reflecting a 67·2% (63·9–70·1%) reduction after seasonal adjustment. Annualized incidence rates of syphilis diagnoses declined 49·0% (42·5–54·7%) over the same period, from 7·7 to 3·8 cases per 100 person-years at risk. In contrast, rates of gonorrhea diagnoses declined only 12·7% (5·7–19·1%), from 13·4 to 11·8 cases per 100 person-years at risk.

**Figure 4:**
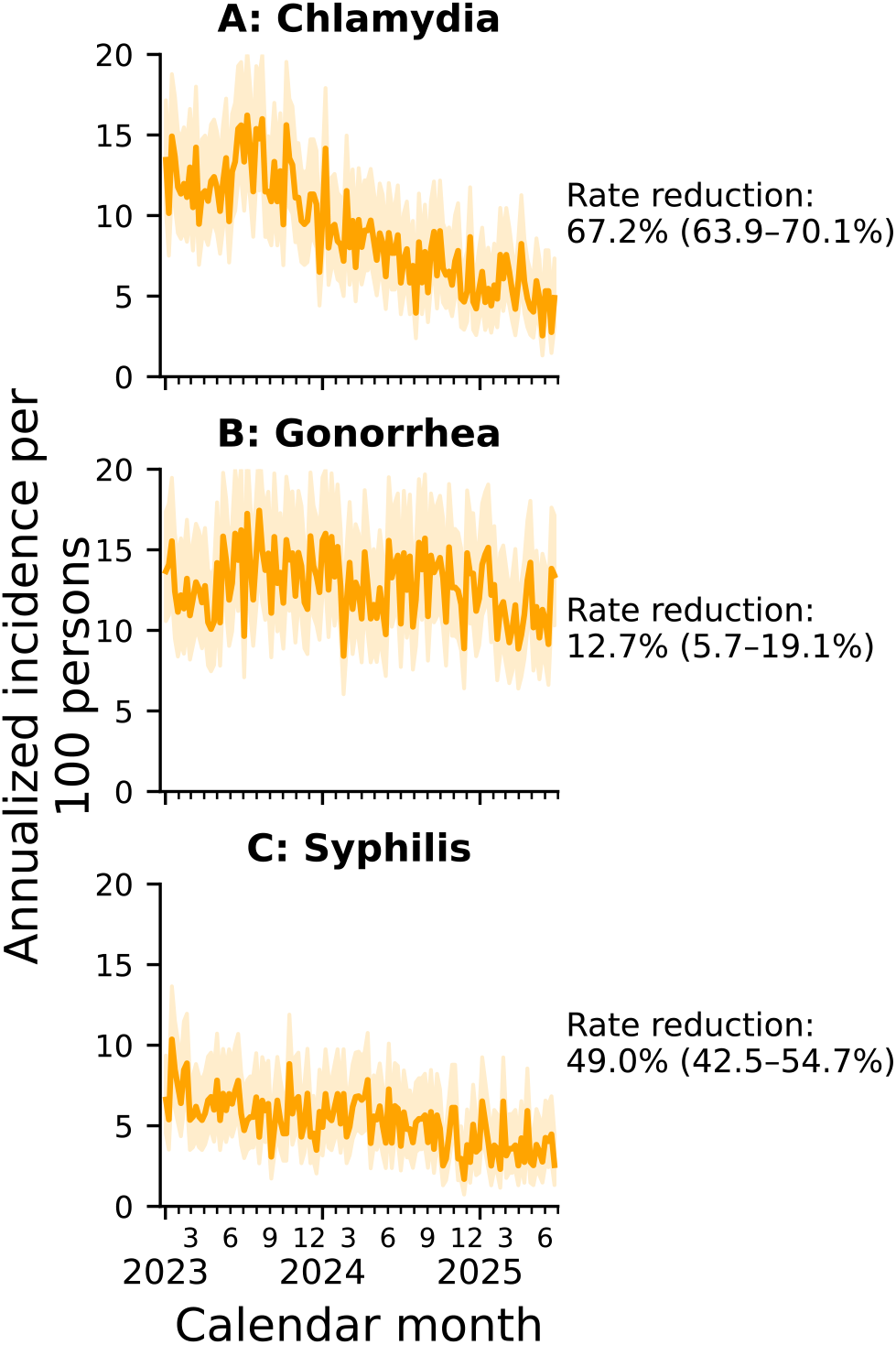
Reduction in incidence of chlamydia, gonorrhea, and syphilis within the study cohort. We illustrate annualized (laboratory-confirmed infections per 100 persons-per year) incidence rates of (**a**) chlamydia, (**b**) gonorrhea, and (**c**) syphilis within the full eligible study cohort, by week throughout the study period. We report accompanying rate reductions comparing annualized incidence rates for the week ending June 30, 2025 to the week beginning January 1, 2023, estimated via Poisson regression models including sine- and cosine transformations of calendar date at 1-year periodicity to adjust for seasonal trends.

## DISCUSSION

We report a rapid loss of effectiveness of doxyPEP for prevention of gonorrhea among males receiving HIV PrEP or living with HIV in a large cohort in southern California. These reductions in effectiveness aligned closely with expansion in prevalence of the *tetM* gene among circulating *N. gonorrhoeae* lineages within the study region. Whereas doxyPEP effectiveness against gonorrhea within 90 days after prescription fills was 47·2% in periods when *tetM* prevalence among circulating *N*. gonorrhoeae was 20-20·9%, effectiveness declined to 35.5%, 21.1% and –8.5% as *tetM* prevalence increased to 30-30·9%, 40-40·9%, and ≥50%, respectively. Within the same cohort, effectiveness of doxyPEP against chlamydia and syphilis was 66·5% and 60·7%, respectively, within 90 days after prescription fills, resembling efficacy against the same infections reported in randomized trials undertaken in the US^6^ and France.^4,5^

Previous US studies undertaken shortly after doxyPEP implementation have reported expansion of *N. gonorrhoeae* lineages harboring *tetM* and other genetic determinants of tetracycline resistance, most notably in West Coast regions where doxyPEP was implemented in advance of US nationwide guidance.^2,3,9,18^ Our findings suggest this expansion has been associated with loss of real-world clinical protection against gonorrhea among doxyPEP recipients. Before doxyPEP implementation, modeling studies employing conservative assumptions estimated that adoption by 10% eligible individuals within a locality would lead to loss of doxycycline effectiveness against gonorrhea within 12·1 years, versus 1·6 years with adoption by 90% of eligible individuals.^10^ Cumulatively over the study period, 8·4% of individuals in our cohort received ≥1 doxyPEP fill, and no more than 4% of individuals had received a prescription fill within the preceding 90 days at any point in our study period. Despite this modest uptake, we estimated that doxyPEP effectiveness against gonorrhea reached 0% between March and June, 2024. This timeframe extends 11-14 months after statewide implementation of doxyPEP in California,^3^ suggesting rapid responses of *N. gonorrhoeae* to doxyPEP-associated selective pressure.^10^

Our study does not provide a direct basis for causally attributing resistance emergence in *N. gonorrhoeae* to doxyPEP implementation. However, several observations support this possibility. First, the timing of observed increases in *tetM* prevalence among locally-circulating *N. gonorrhoeae* aligns closely with statewide doxyPEP implementation in California. Second, cohort-wide reductions in incidence of chlamydia (66·3%) and syphilis (49·2%), despite modest doxyPEP uptake, substantiate the notion that doxyPEP recipients wield outsized influence on broader STI transmission dynamics.^19^ Enhanced risk of infection with tetracycline-resistant *N. gonorrhoeae* lineages among a “core group” of doxyPEP recipients—as documented in France^8^ and Seattle^20^—may thus shape antimicrobial susceptibility of *N. gonorrhoeae* at a broader population level. Additional studies document associations of antimicrobial resistance with doxyPEP receipt in other bacterial species, including group A *Streptococcus* and *Staphylococcus aureus*,^20^ non-gonorrhea *Neisseria* species,^21^ and gut microbiome commensals.^22^ Observations from our study mirror historical experience with minocycline prophylaxis for gonorrhea, which was found to offer limited value due to rapid resistance selection.^23^

Strengths of our study include the use of a test-negative design to mitigate confounding driven by healthcare-seeking behavior, our ability to control for detailed aspects of individuals’ STI risk based on demographic factors and clinical history, and the opportunity to relate doxyPEP effectiveness to trends in *tetM* prevalence among *N. gonorrhoeae* within the study region. Several limitations of our study also merit consideration. First, confounding due to unmeasured variables remains possible under our observational study design. However, our findings that doxyPEP was effective against chlamydia and syphilis—and against gonorrhea during periods when *tetM* prevalence was low in *N. gonorrhoeae*—closely align with findings from pre-implementation randomized trials.^4–6^ Second, our study did not measure the frequency and quantity of doxycycline consumption among doxyPEP recipients. Our finding that effectiveness was greatest within the period shortly following prescription fills nonetheless supports the biological plausibility of our estimates. We observe considerable variation by time since individuals’ last doxyPEP fill in treatment effectiveness against chlamydia (diagnosed by molecular testing), and only modest differences in effectiveness against syphilis (diagnosed by serological testing).

Third, we do not characterize resistance individually among cohort members who received or did not receive doxyPEP, nor do we substantiate whether resistance was a factor in treatment failure for individuals. For at least two reasons, regional *tetM* prevalence data in our study likely underestimated the proportion of gonococcal infections expected to show diminished susceptibility to doxycycline. Although the association of *tetM* with high-level tetracycline resistance^7^ makes this gene a useful target for surveillance, genetic factors besides *tetM* impact tetracycline susceptibility^24^ and are not addressed in our analyses. Moreover, gonococcal isolates from MSM are more likely than those from non-MSM to exhibit AMR,^25^ and MSM account for only ~32% of GISP isolates.^26^ Last, our study does not quantify real-world uptake of doxyPEP. While KPSC recipients closely resemble the regional population in demographic and other characteristics,^12^ clinical practices within the healthcare system may not be generalizable to other providers.

These findings may inform benefit and risk considerations for ongoing doxyPEP implementation. Prior to doxyPEP implementation, incidence of STIs showed increasing trends in numerous settings,^27^ and growing proportions of MSM reported condomless sex with casual partners.^28,29^ While our analyses and others^11,30–32^ demonstrate real-world effectiveness of doxyPEP against chlamydia and syphilis, gonorrhea accounts for many STI episodes initially expected to be preventable by doxyPEP,^33^ and a substantial share of all preventable morbidity due to the associated risk for complications including epididymitis, orchitis, and disseminated gonococcal infection.^34^ Rapid resistance emergence in *N. gonorrhoeae* may undermine this benefit, particularly as *tetM* frequently co-occurs with genetic determinants of resistance to therapeutically-relevant drug classes including cephalosporins.^35,36^ Implications of potential resistance selection in other pathogens remain to be understood.^37^ Accelerated implementation of doxyPEP in West Coast regions of the US, and associated expansion of lineages harboring *tetM*,^9^ provided an early opportunity to observe changes in effectiveness of this intervention against gonorrhea in our study cohort. Monitoring for similar changes through ongoing surveillance remains an important objective amid broader doxyPEP implementation in other geographic settings.

## Data Availability

The raw data used in this manuscript include protected health information (e.g., dates of diagnoses and testing) that cannot be shared openly without appropriate ethical oversight and approval as well as institutional data use agreements. Kaiser Permanente Southern California (KPSC) institutional policy requires a data transfer agreement to be executed between KPSC and the individual recipient entity prior to transmittal of patient-level data outside KPSC. Requests for data can be addressed to.

## ACKNOWLEDGMENTS

This work was supported by the Center for Forecasting and Outbreak Analytics of the US Centers for Disease Control & Prevention (CDC) via the Insight Net cooperative agreement (CDC-RFA-FT-23-0069 to SYT and JAL). Its contents are solely the responsibility of the authors and do not necessarily represent the official views of the Centers for Disease Control and Prevention. The authors thank the CDC and CDC-funded public health programs for their work collecting and characterizing *Neisseria gonorrhoeae* isolates and for making resulting *N. gonorrhoeae* genome sequences publicly available via the National Center for Biotechnology Information (NCBI) GenBank database.

