## Supporting information for "Rapid loss of doxycycline effectiveness against gonorrhea after implementation of post-exposure prophylaxis in southern California: an observational study"

* These authors contributed equally

**Contents of this supplement**


| Item | Title | Page |
|---|---|---|
| Text S1 | Genomic analyses. | 2 |
| Text S2 | Logistic regression models and time to loss of effectiveness analyses. | 3 |
| Text S3 | Supplementary references. | 5 |
| Table S1 | Sexually-transmitted infection diagnosis codes. | 6 |
| Table S2 | Classification of prescription medications by therapeutic group. | 7 |
| Table S3 | Characteristics of the study cohort. | 8 |
| Table S4 | Effectiveness of doxyPEP against chlamydia, gonorrhea, and syphilis in analyses limited to individuals with recent STI diagnoses. | 9 |
| Table S5 | Effectiveness of doxyPEP against chlamydia, gonorrhea, and syphilis in analyses limited to individuals with recent gonorrhea testing. | 10 |
| Table S6 | Prevalence of *tetM* among sequenced gonococcal isolates in the study region, by month and isolates source. | 11 |
| Table S7 | Effectiveness of doxyPEP against gonorrhea, by *tetM* prevalence (derived from all *N. gonorrhoeae* isolates). | 12 |
| Table S8 | Comparison of model fit with differing continuous transformations of effect-modifying covariates. | 13 |
| Figure S1 | Prevalence of *tetM* among sequenced gonococcal isolates in the study region, with observed estimates and weighted LOESS smoothing. | 14 |
| Figure S2 | Variation in doxyPEP effectiveness with *tetM* prevalence among all gonococcal isolates, and time to loss of effect. | 15 |

**Text S1: Genomic analyses.**

We obtained sequencing data for isolates collected at sentinel sites within Los Angeles, Orange and San Diego counties, which were collected at clinics participating in the US Centers for Disease Control and Prevention (CDC) Gonococcal Isolate Surveillance Project (GISP) between the years 2022–2024.[1] Isolates submitted for inclusion in GISP are collected from the first 25 men each month with symptomatic gonococcal urethritis who receive care at participating sexually-transmitted disease clinics in each sentinel region. Additional sensitivity analyses included further isolates collected as part of two expanded CDC-led surveillance programs operated within the same sentinel clinics: the Enhanced Gonococcal Isolate Surveillance Program (eGISP) and Strengthening the US Response to Resistant Gonorrhea (SURRG) program,[2] which further includes extragenital *N. gonorrhoeae* isolates and cervical or vaginal gonococcal isolates obtained from women. National Center for Biotechnology Information (NCBI) GenBank accession numbers of the isolates included in the study are listed in **Sheet S1**.

De novo assembly was performed using SPAdes[3] (v 3.12.0) using the –careful flag. Reference-based mapping to NCCP11945 (NC_011035.1) was performed using BWA-MEM[4] (v 0.7.17). We used Pilon[5] (v 1.23) to call variants (minimum mapping quality: 20, minimum coverage: 10X) after marking duplicate reads with Picard v 2.20.1 (https://broadinstitute.github.io/picard/) and sorting reads using samtools[6] (v 1.17). We generated pseudogenomes by incorporating variants supported by at least 90 % of reads and sites with ambiguous alleles into the reference genome sequence. We mapped reads to a single copy of the locus encoding the 23S rRNA and called variants using the same procedures.[7]

To determine the presence or absence of *tetM*, we used the results of blastn[8] (v 2.9.0) searches of de novo assemblies for *tetM* (MG874353.1). We determined that *tetM* was present if the blast hit was >50% of the query length with >95% identity.

We filtered assembled genomes based on the following criteria: (1) The total assembly length was longer than 1,900,000 bp and less than 2,300,000 bp; (2) Reference coverage was more than 30%; (3) Percentage of reads mapped to reference was at least 70%; (4) Less than 12% of positions were missing in pseudogenomes.

**Text S2: Logistic regression models and time to loss of effectiveness analyses.**

*Continuous-modifier models*

To evaluate effect modification in the association between receipt of doxyPEP and detection of *N. gonorrhoeae*, we fit logistic regression models defining interaction terms between exposure (doxyPEP fill within 90 days before testing) and either calendar time or the prevalence of *tetM*. Both modifiers were treated as continuous variables. Calendar time was represented by a numeric index indicating the calendar month of the test. To estimate *tetM* prevalence, we applied weighted LOESS smoothing, with weights proportional to the total number of isolates collected in the study region each month (**Figure S1; Table S6**).

For months without isolate collection, prevalence values were projected using the LOESS fit. Separate models were constructed for GISP isolates alone and for the combined dataset including both GISP and non-GISP isolates.

All regression models were adjusted for the following covariates: age group (16–25, 26–35, 36–45, ≥46 years); race/ethnicity (Hispanic, White non-Hispanic, Black non-Hispanic, Asian/Pacific Islander non-Hispanic, Other/Unknown); gender identity (cisgender or transgender, ascertained using natural language processing of structured and unstructured HER fields[9]); HIV infection or receipt of HIV PrEP/PEP in the prior 180 days; prior-year number of gonorrhea tests (continuous); prior gonorrhea and chlamydia diagnoses (0, 1, ≥2); history of syphilis (yes/no); history of JYNNEOS vaccination (yes/no); commercial insurance (yes/no); community-level socioeconomic status using the neighborhood deprivation index (continuous); and antibiotic use in the preceding 30 days (no treatment, STI-related treatment, treatment unrelated to STI; **Table S2**).

For models examining calendar time as an effect modifier, we additionally adjusted for seasonality using sine and cosine transformations of calendar date assuming 12-month periodicity.

*Model selection*

To avoid imposing *a priori* assumptions about the functional form of calendar time or *tetM* prevalence, we evaluated several transformations for each modifier, including linear, square-root, quadratic, cubic, and logarithmic functions; for *tetM* prevalence we additionally assessed a logit transformation. Model performance was compared using the area under the receiver operating characteristic curve (AUC), Akaike Information Criterion (AIC), and Bayesian Information Criterion (BIC; **Table S8**).

For both time and *tetM* modifiers, the linear specification yielded comparable or lower AIC and BIC values than all alternative transformations, while AUC was similar to the quadratic and cubic forms. Based on these criteria, we selected a linear transformation for both modifiers.

*Time to loss of doxyPEP effectiveness*

To quantify the time at which doxyPEP fill withing the preceding 90 days ceased to be protective against *N. gonorrhoeae* infection, we first computed adjusted odds ratios (aORs) across calendar time and across *tetM* prevalence using the fitted interaction models.

For the time-interaction model, the aOR is:

$$\text{aOR}(t) = \exp\left(\beta_x + \beta_{x:T} \times t\right) \quad (1)$$

where *t* is the calendar time, $\beta_x$ is the coefficient for having received a doxyPEP fill in the prior 90 days, and $\beta_{x:T}$ is the coefficient for the interaction between doxyPEP use and calendar time.

Effectiveness is lost when:

$$\text{aOR} = 1 \Leftrightarrow \ln\left(\text{aOR}\right) = 0. \quad (2)$$

We define the time for loss of protection as *t**, such that

$$t^* = -\frac{\beta_x}{\beta_{x:T}} \quad (3)$$

which represents the time index or *tetM* prevalence value at which the estimated protective association disappears.

To project time to loss of protection based on *tetM* prevalence, we defined

(4) $$\mathrm{aOR}(p) = \exp\left(\beta_x + \beta_{x:P} \times p\right)$$

where *p* indicated the smoothed monthly prevalence of *tetM* (based on the LOESS fit). Defining $p^* = -\beta_x/\beta_{x:P}$, similar to Equation (3), we defined time to loss of protection as the first month in which the smoothed monthly *tetM* prevalence (based on the LOESS fit) exceeded *p** (**Figure S1**).

*Uncertainty estimation*

To characterize the distribution of $t^*$ and $p^*$ values, we performed a bootstrap procedure with 100,000 iterations. In each iteration, we sampled the coefficient vectors $(\beta_x, \beta_{x:T};\ \beta_x, \beta_{x:P})$ from a multivariate normal distribution defined by each model's estimated coefficients and their covariance matrix. To map *tetM* prevalence to calendar time, each bootstrap iteration generated a monthly *tetM* prevalence trajectory by drawing monthly *tetM* counts from a multinomial distribution ($\vec{p}$ =observed monthly *tetM* prevalence and $\vec{N}$ =total number of *N. gonorrhoeae* isolates collected each month). The resulting resampled monthly prevalence series was then smoothed using weighted LOESS, with weights proportional to the number of isolates sequenced per month, to obtain a continuous calendar-time trajectory. For months without isolate collection, prevalence values were interpolated using the LOESS fit. The resulting distributions were used directly to construct the boxplots.

**Text S3: Supplementary references.**


1 Pham C, Raphael B, St. Cyr S, Torrone E. Gonococcal Isolate Surveillance Project (GISP) and Enhanced GISP (eGISP). 2020. http://stacks.cdc.gov/view/cdc/108436 (accessed Dec 19, 2025).

2 Schlanger K, Learner ER, Pham CD, *et al.* Strengthening the US Response to Resistant Gonorrhea: an overview of a multisite program to enhance local response capacity for antibiotic-resistant *Neisseria gonorrhoeae*. *Sex Transm Dis* 2021; 48: S97–103.

3 Bankevich A, Nurk S, Antipov D, *et al.* SPAdes: a new genome assembly algorithm and its applications to single-cell sequencing. *J Comput Biol* 2012; 19: 455–77.

4 Li H. Aligning sequence reads, clone sequences and assembly contigs with BWA-MEM. 2013; published online March 16. https://arxiv.org/pdf/1303.3997 (accessed Dec 19, 2025).

5 Walker BJ, Abeel T, Shea T, *et al.* Pilon: An Integrated Tool for Comprehensive Microbial Variant Detection and Genome Assembly Improvement. *PLoS One* 2014; 9: e112963.

6 Li H, Handsaker B, Wysoker A, *et al.* The Sequence Alignment/Map format and SAMtools. *Bioinformatics* 2009; 25: 2078–9.

7 Johnson SR, Grad Y, Abrams AJ, Pettus K, Trees DL. Use of whole-genome sequencing data to analyze 23S rRNA-mediated azithromycin resistance. *Int J Antimicrob Agents* 2017; 49: 252–4.

8 Camacho C, Coulouris G, Avagyan V, *et al.* BLAST+: architecture and applications. *BMC Bioinformatics* 2009; 10. DOI:10.1186/1471-2105-10-421.

9 Xie F, Getahun D, Quinn VP, *et al.* An automated algorithm using free-text clinical notes to improve identification of transgender people. *Inform Health Soc Care* 2021; 46: 18–28.

**Table S1: Sexually-transmitted infection diagnosis codes.**

| Diagnosis | ICD-10 code |
|---|---|
| HIV | |
| | B20 |
| | B97.35 |
| | O98.711 |
| | O98.712 |
| | O98.713 |
| | O98.719 |
| | O98.72 |
| | O98.73 |
| | R75 |
| | Z21 |
| Mpox | |
| | B04 |
| Body lice | |
| | B85.1 |
| Anogenital warts | |
| | A63 |
| Canchroid | |
| | A57 |
| Nonspecific urethritis | |
| | N34.1 |
| | N34.2 |
| | N34.3 |
| Gonococcal infection | |
| | A54* |
| Chlamydia infection | |
| | A56* |
| Incident syphilis infection | |
| | A51.0 |
| | A51.1 |
| | A51.2 |
| | A51.5 |
| | A51.9 |
| Substance use disorders | |
| | F10-F19 |

**Table S2: Classification of prescription medications by therapeutic group.**

| Medication | Medication names |
|---|---|
| HIV Pre-/Post-Exposure Prophylaxis (PrEP/PEP) | Emtricitabine Tenofovir Disoproxil Fumarate; Emtricitabine Tenofovir Alafenamide; Efavirenz Emtricitabine Tenofovir DF; Lamivudine Tenofovir Disoproxil Fumarate; Emtricitabine; Tenofovir Alafenamide; Tenofovir Disoproxil Fumarate |
| HIV Antiretroviral Therapy (ART) | Bictegravir Emtricitabine Tenofovir Alafenamide; Emtricitabine Rilpivirine Tenofovir Alafenamide; Elvitegravir Cobicistat Emtricitabine Tenofovir Alafenamide; Elvitegravir Cobicistat Emtricitabine Tenofovir Disoproxil Fumarate; Darunavir Cobicistat Emtricitabine Tenofovir Alafenamide |
| Doxycycline Post-Exposure Prophylaxis (doxyPEP) | Doxycycline Monohydrate; Doxycycline Hyclate |
| Penicillins | Amoxicillin; Ampicillin; Oxacillin; Penicillin |
| β-Lactam / β-Lactamase Inhibitor Combinations | Amoxicillin-Clavulanate; Ampicillin-Sulbactam; Piperacillin-Tazobactam |
| Cephalosporins | Cefazolin; Cefprozil; Cefuroxime; Cefdinir; Ceftriaxone; Cefotaxime; Ceftazidime; Cefepime; Ceftaroline |
| Carbapenems | Meropenem; Ertapenem; Imipenem; Imipenem-Cilastatin |
| Monobactam | Aztreonam |
| Macrolides | Azithromycin; Clarithromycin; Erythromycin |
| Fluoroquinolones | Ciprofloxacin; Levofloxacin; Moxifloxacin; Gemifloxacin |
| Aminoglycosides | Gentamicin; Tobramycin; Amikacin |
| Tetracyclines | Doxycycline; Tetracycline |
| Lincosamide | Clindamycin |
| Glycopeptide | Vancomycin |
| Oxazolidinone | Linezolid |
| Sulfonamide Combination | Trimethoprim-Sulfamethoxazole (TMP-SMX); Trimethoprim; Sulfamethoxazole |
| Rifamycin | Rifampin |

**Table S3: Characteristics of the study cohort.**

| Characteristics at baseline | | Individuals, *n* (%) | |
|---|---|---|---|
| | | DoxyPEP ever received | No doxyPEP receipt |
| | | *N*=2,262 | *N*=24,320 |
| Age group | | | |
| | 16-25 years | 351 (15·5) | 3,040 (12·5) |
| | 26-35 years | 554 (24·5) | 5,063 (20·8) |
| | 36-45 years | 896 (39·6) | 7,631 (31·4) |
| | ≥46 years | 461 (20·4) | 8,586 (35·3) |
| | Median (interquartile range) | 34 (29–43) | 38 (30–53) |
| Race/ethnicity | | | |
| | White, non-Hispanic | 592 (26·2) | 7,962 (32·7) |
| | Black, non-Hispanic | 159 (7·0) | 2,246 (9·2) |
| | Hispanic, any race | 1,102 (48·7) | 9,932 (40·8) |
| | Asian/Pacific Islander, non-Hispanic | 229 (10·1) | 2,202 (9·1) |
| | Other/Unknown | 180 (8·0) | 1,978 (8·1) |
| Gender identity | | | |
| | No evidence of transgender identity | 2,217 (98·0) | 23,949 (98·5) |
| | Transgender identity recorded | 45 (2·0) | 371 (1·5) |
| Health insurance source | | | |
| | Commercial plan | 1,982 (87·6) | 20,463 (84·1) |
| | Others/Unknown | 280 (12·4) | 3,857 (15·9) |
| Neighborhood deprivation index | | | |
| | Quartile 1 (least deprived) | 554 (24·5) | 6,093 (25·1) |
| | Quartile 2 | 510 (22·5) | 6,142 (25·3) |
| | Quartile 3 | 585 (25·9) | 6,057 (24·9) |
| | Quartile 4 (most deprived) | 613 (27·1) | 6,028 (24·8) |
| HIV infection status or prevention | | | |
| | HIV infection | 157 (6·9) | 4,005 (16·5) |
| | HIV pre/post-exposure prophylaxis received | 913 (40·4) | 7,482 (30·8) |
| Mpox vaccination status | | | |
| | 0 JYNNEOS doses | 684 (30·2) | 11,859 (48·8) |
| | ≥1 JYNNEOS dose | 1,578 (69·8) | 12,461 (51·2) |
| Prior-year STI tests | | | |
| | 0-2 tests | 1,086 (48·0) | 14,972 (61·6) |
| | 3-5 tests | 861 (38·1) | 7,750 (31·9) |
| | 6-11 tests | 297 (13·1) | 1,548 (6·4) |
| | ≥12 tests | 18 (0·8) | 50 (0·2) |
| Prior chlamydia diagnoses | | | |
| | 0 diagnoses | 1,955 (86·4) | 22,252 (91·5) |
| | 1 diagnosis | 248 (11·0) | 1,778 (7·3) |
| | ≥2 diagnoses | 59 (2·6) | 290 (1·2) |
| Prior gonorrhea diagnoses | | | |
| | 0 diagnoses | 1,906 (84·3) | 22,097 (90·9) |
| | 1 diagnosis | 272 (12·0) | 1,857 (7·6) |
| | ≥2 diagnoses | 84 (3·7) | 366 (1·5) |
| Prior syphilis diagnosis | | | |
| | No | 1,846 (81·6) | 20,555 (84·5) |
| | Yes | 416 (18·4) | 3,765 (15·5) |
| Recent (30d) antibiotic treatment | | | |
| | No treatment | 1,990 (88·0) | 22,501 (92·5) |
| | STI-related treatment | 73 (3·2) | 343 (1·4) |
| | Treatment unrelated to STI | 199 (8·8) | 1,476 (6·1) |
| New diagnosis in prior year | | | |
| | No | 489 (21·6) | 5,989 (24·6) |
| | Yes | 1,773 (78·4) | 18,331 (75·4) |

### Table S4: Effectiveness of doxyPEP against chlamydia, gonorrhea, and syphilis in analyses limited to individuals with recent STI diagnoses.

| Subgroup | Outcome | Most recent doxyPEP fill | Tests, *n* (%) | | Odds ratio (95% confidence interval) | | Treatment effectiveness (95% confidence interval) |
|---|---|---|---|---|---|---|---|
| | | | Negative | Positive | Unadjusted | Adjusted | Adjusted |
| Individuals with ≥1 STI diagnosis in prior 12 months | | | | | | | |
| | Chlamydia | | | | | | |
| | | | *N*=32,414 | *N*=2,131 | | | |
| | | Never received | 29,456 (90·9%) | 2,044 (95·9%) | ref. | ref. | ref. |
| | | >180d before testing | 1,038 (3·2%) | 47 (2·2%) | 0·65 (0·47–0·90) | 0·87 (0·64–1·19) | 12·6% (-18·7–35·7) |
| | | 91–180d before testing | 688 (2·1%) | 14 (0·7%) | 0·29 (0·17–0·49) | 0·38 (0·23–0·65) | 61·6% (34·7–77·5) |
| | | <90d before testing | 1,232 (3·8%) | 26 (1·2%) | 0·30 (0·20–0·45) | 0·42 (0·28–0·63) | 58·2% (37·0–72·2) |
| | Gonorrhea | | *N* =31,450 | *N* =3,078 | | | |
| | | Never received | 28,687 (91·2%) | 2,798 (90·9%) | ref. | ref. | ref. |
| | | >180d before testing | 986 (3·1%) | 99 (3·2%) | 1·03 (0·84–1·27) | 1·02 (0·83–1·25) | -2·0% (-25·2–16·8) |
| | | 91–180d before testing | 643 (2·0%) | 60 (1·9%) | 0·96 (0·72–1·27) | 0·98 (0·74–1·29) | 2·4% (-28·9–26·1) |
| | | <90d before testing | 1,134 (3·6%) | 121 (3·9%) | 1·09 (0·89–1·34) | 1·05 (0·86–1·29) | -5·5% (-28·9–13·7) |
| | Syphilis | | *N* =8,636 | *N* =287 | | | |
| | | Never received | 7,751 (89·8%) | 273 (95·1%) | ref. | ref. | ref. |
| | | >180d before testing | 318 (3·7%) | 8 (2·8%) | 0·71 (0·36–1·44) | 0·97 (0·48–1·96) | 2·8% (-96·0–51·8) |
| | | 91–180d before testing | 212 (2·5%) | 4 (1·4%) | 0·54 (0·20–1·43) | 0·62 (0·22–1·73) | 38·1% (-73·1–77·8) |
| | | <90d before testing | 355 (4·1%) | 2 (0·7%) | 0·16 (0·04–0·65) | 0·24 (0·06–0·99) | 76·3% (1·3–94·3) |
| Individuals with ≥1 STI diagnosis in prior 6 months | | | | | | | |
| | Chlamydia | | | | | | |
| | | | *N*=17,051 | *N*=1,197 | | | |
| | | Never received | 15,479 (90·8%) | 1,147 (95·8%) | ref. | ref. | ref. |
| | | >180d before testing | 493 (2·9%) | 25 (2·1%) | 0·68 (0·44–1·06) | 0·95 (0·62–1·46) | 4·7% (-45·5–37·5) |
| | | 91–180d before testing | 363 (2·1%) | 9 (0·8%) | 0·34 (0·17–0·64) | 0·46 (0·24–0·90) | 53·8% (10·1–76·2) |
| | | <90d before testing | 716 (4·2%) | 16 (1·3%) | 0·30 (0·18–0·51) | 0·43 (0·25–0·73) | 57·0% (27·0–74·6) |
| | Gonorrhea | | *N*=16,513 | *N*=1,719 | | | |
| | | Never received | 15,033 (91·0%) | 1,580 (91·9%) | ref. | ref. | ref. |
| | | >180d before testing | 478 (2·9%) | 40 (2·3%) | 0·80 (0·58–1·10) | 0·81 (0·59–1·13) | 18·7% (-12·6–41·2) |
| | | 91–180d before testing | 338 (2·0%) | 34 (2·0%) | 0·96 (0·67–1·38) | 1·04 (0·72–1·50) | -4·1% (-50·2–27·9) |
| | | <90d before testing | 664 (4·0%) | 65 (3·8%) | 0·93 (0·70–1·25) | 0·96 (0·73–1·27) | 3·7% (-27·5–27·2) |
| | Syphilis | | *N*=4,332 | *N*=135 | | | |
| | | Never received | 3,872 (89·4%) | 129 (95·6%) | ref. | ref. | ref. |
| | | >180d before testing | 146 (3·4%) | 2 (1·5%) | 0·41 (0·10–1·71) | 0·62 (0·15–2·61) | 37·6% (-161·1–85·1) |
| | | 91–180d before testing | 111 (2·6%) | 3 (2·2%) | 0·81 (0·26–2·56) | 1·06 (0·30–3·69) | -6·1% (-269·3–69·5) |
| | | <90d before testing | 203 (4·7%) | 1 (0·7%) | 0·15 (0·02–1·06) | 0·24 (0·03–1·83) | 75·5% (-82·6–96·7) |

We estimated adjusted odds ratios via logistic regression models controlling for: age group; race/ethnicity; transgender identity; commercial or non-commercial insurance source; neighborhood deprivation index; HIV infection status or PrEP receipt; prior receipt of JYNNEOS; prior-year gonorrhea test frequency and history of diagnoses with chlamydia, gonorrhea, or syphilis; and recent (30d) antibiotic receipt. Results of all tests were included, provided individuals did not receive a positive result for the studied pathogen within the preceding 30 days for chlamydia and gonorrhea and 365 days for syphilis. We define treatment effectiveness as $(1 - \mathrm{aOR}) \times 100\%$. Analyses of individuals with ≥1 STI diagnosis in the prior 12 months included 9,366 unique individuals tested for chlamydia, 9,363 tested for gonorrhea, and 2,785 tested for syphilis. Analyses of individuals with ≥1 STI diagnosis in the prior 6 months included 6,941 unique individuals tested for chlamydia, 6,936 tested for gonorrhea, and 1,892 tested for syphilis. Results of all tests were included, provided individuals did not receive a positive result for the studied pathogen within the preceding 30 days for chlamydia and gonorrhea and 365 days for syphilis. We define treatment effectiveness as $(1 - \mathrm{aOR}) \times 100\%$.

**Table S5: Effectiveness of doxyPEP against chlamydia, gonorrhea, and syphilis in analyses limited to individuals with recent gonorrhea testing.**

| Subgroup | Outcome | Most recent doxyPEP fill | Tests, *n* (%) | | Odds ratio (95% confidence interval) | | Treatment effectiveness (95% confidence interval) |
|---|---|---|---|---|---|---|---|
| | | | Negative | Positive | Unadjusted | Adjusted | Adjusted |
| Individuals with ≥4 gonorrhea tests in prior 12 months | | | | | | | |
| | Chlamydia | | | | | | |
| | | | *N*=52,339 | *N*=2,211 | | | |
| | | Never received | 47,146 (90·1·0%) | 2,099 (94·9·0%) | ref. | ref. | ref. |
| | | >180d before testing | 1,960 (3·7·0%) | 65 (2·9·0%) | 0·74 (0·57–0·98) | 0·98 (0·76–1·26) | 2·1% (-26·3–24·1) |
| | | 91–180d before testing | 1,144 (2·2·0%) | 20 (0·9·0%) | 0·39 (0·25–0·61) | 0·48 (0·31–0·75) | 52·1% (25·1–69·3) |
| | | <90d before testing | 2,089 (4·0·0%) | 27 (1·2·0%) | 0·29 (0·20–0·43) | 0·36 (0·24–0·53) | 64·1% (46·6–75·9) |
| | Gonorrhea | | *N*=51,086 | *N*=3,470 | | | |
| | | Never received | 46,153 (90·3·0%) | 3,099 (89·3·0%) | ref. | ref. | ref. |
| | | >180d before testing | 1,898 (3·7·0%) | 127 (3·7·0%) | 1·00 (0·82–1·21) | 0·99 (0·82–1·19) | 1·4% (-18·8–18·1) |
| | | 91–180d before testing | 1,081 (2·1·0%) | 86 (2·5·0%) | 1·19 (0·95–1·48) | 1·16 (0·93–1·46) | -16·1% (-45·5–7·3) |
| | | <90d before testing | 1,954 (3·8·0%) | 158 (4·6·0%) | 1·20 (1·00–1·44) | 1·08 (0·91–1·28) | -7·9% (-28·5–9·4) |
| | Syphilis | | *N*=11,437 | *N*=295 | | | |
| | | Never received | 10,123 (88·5·0%) | 276 (93·6·0%) | ref. | ref. | ref. |
| | | >180d before testing | 461 (4·0·0%) | 9 (3·1·0%) | 0·72 (0·37–1·44) | 0·94 (0·48–1·85) | 5·8% (-85·2–52·1) |
| | | 91–180d before testing | 296 (2·6·0%) | 7 (2·4·0%) | 0·87 (0·41–1·84) | 1·08 (0·49–2·37) | -7·6% (-136·9–51·1) |
| | | <90d before testing | 557 (4·9·0%) | 3 (1·0·0%) | 0·20 (0·06–0·62) | 0·27 (0·09–0·88) | 72·6% (11·7–91·5) |
| Individuals with ≥2 gonorrhea tests in prior 12 months | | | | | | | |
| | Chlamydia | | | | | | |
| | | | *N*=99,534 | *N*=3,713 | | | |
| | | Never received | 91,553 (92·0%) | 3,557 (95·8%) | ref. | ref. | ref. |
| | | >180d before testing | 3,191 (3·2%) | 95 (2·6%) | 0·77 (0·61–0·96) | 0·91 (0·73–1·12) | 9·5% (-12·0–26·9) |
| | | 91–180d before testing | 1,788 (1·8%) | 25 (0·7%) | 0·36 (0·24–0·53) | 0·40 (0·27–0·60) | 59·7% (40·0–73·0) |
| | | <90d before testing | 3,002 (3·0%) | 36 (1·0%) | 0·31 (0·22–0·43) | 0·35 (0·25–0·49) | 65·1% (50·9–75·2) |
| | Gonorrhea | | *N*=97,696 | *N*=5,532 | | | |
| | | Never received | 90,070 (92·2%) | 5,025 (90·8%) | ref. | ref. | ref. |
| | | >180d before testing | 3,087 (3·2%) | 197 (3·6%) | 1·14 (0·98–1·33) | 1·04 (0·90–1·21) | -4·4% (-20·8–9·7) |
| | | 91–180d before testing | 1,701 (1·7%) | 114 (2·1%) | 1·20 (0·99–1·46) | 1·10 (0·91–1·34) | -10·1% (-34·0–9·5) |
| | | <90d before testing | 2,838 (2·9%) | 196 (3·5%) | 1·24 (1·05–1·46) | 1·00 (0·86–1·18) | -0·4% (-17·6–14·3) |
| | Syphilis | | *N*=20,614 | *N*=691 | | | |
| | | Never received | 18,689 (90·7%) | 662 (95·8%) | ref. | ref. | ref. |
| | | >180d before testing | 741 (3·6%) | 15 (2·2%) | 0·57 (0·34–0·96) | 0·72 (0·43–1·21) | 27·9% (-20·9–57·0) |
| | | 91–180d before testing | 426 (2·1%) | 8 (1·2%) | 0·53 (0·26–1·07) | 0·66 (0·33–1·36) | 33·6% (-35·9–67·5) |
| | | <90d before testing | 758 (3·7%) | 6 (0·9%) | 0·22 (0·10–0·50) | 0·32 (0·14–0·72) | 68·3% (28·0–86·1) |

We estimated adjusted odds ratios via logistic regression models controlling for: age group; race/ethnicity; transgender identity; commercial or non-commercial insurance source; neighborhood deprivation index; HIV infection status or PrEP receipt; prior receipt of JYNNEOS; prior-year gonorrhea test frequency and history of diagnoses with chlamydia, gonorrhea, or syphilis; and recent (30d) antibiotic receipt. Results of all tests were included, provided individuals did not receive a positive result for the studied pathogen within the preceding 30 days for chlamydia and gonorrhea and 365 days for syphilis. We define treatment effectiveness as $(1 - \mathrm{aOR}) \times 100\%$. Analyses of individuals with ≥4 tests in the prior year included 10,560 unique individuals tested for chlamydia, 10,561 tested for gonorrhea, and 2,612 tested for syphilis. Analyses of individuals with ≥2 tests in the prior year included 19,273 unique individuals tested for chlamydia, 19,273 tested for gonorrhea, and 4,765 tested for syphilis. Results of all tests were included, provided individuals did not receive a positive result for the studied pathogen within the preceding 30 days for chlamydia and gonorrhea and 365 days for syphilis. We define treatment effectiveness as $(1 - \mathrm{aOR}) \times 100\%$.

**Table S6: Prevalence of *tetM* among sequenced gonococcal isolates in the study region, by month and isolates source.**

| Month | GISP isolates | | All isolates | |
|---|---|---|---|---|
| | Count, *tetM* present/total sequenced | Proportion, % | Count, *tetM* present/total sequenced | Proportion, % |
| January, 2022 | 5 / 18 | 27·78 | 15 / 52 | 28·85 |
| February, 2022 | 1 / 17 | 5·88 | 5 / 30 | 16·67 |
| March, 2022 | 1 / 14 | 7·14 | 2 / 32 | 6·25 |
| April, 2022 | 3 / 15 | 20·00 | 11 / 43 | 25·58 |
| May, 2022 | 4 / 16 | 25·00 | 10 / 41 | 24·39 |
| June, 2022 | 0 / 19 | 0·00 | 5 / 40 | 12·50 |
| July, 2022 | 2 / 14 | 14·29 | 3 / 26 | 11·54 |
| August, 2022 | 7 / 9 | 77·78 | 13 / 24 | 54·17 |
| September, 2022 | 6 / 13 | 46·15 | 11 / 26 | 42·31 |
| October, 2022 | 4 / 15 | 26·67 | 7 / 20 | 35·00 |
| November, 2022 | 3 / 11 | 27·27 | 7 / 27 | 25·93 |
| December, 2022 | 3 / 14 | 21·43 | 3 / 17 | 17·65 |
| January, 2023 | 2 / 14 | 14·29 | 4 / 30 | 13·33 |
| February, 2023 | 4 / 19 | 21·05 | 5 / 25 | 20·00 |
| March, 2023 | 5 / 19 | 26·32 | 12 / 41 | 29·27 |
| April, 2023 | 4 / 17 | 23·53 | 8 / 36 | 22·22 |
| May, 2023 | 9 / 16 | 56·25 | 18 / 37 | 48·65 |
| June, 2023 | 1 / 15 | 6·67 | 3 / 38 | 7·89 |
| July, 2023 | 7 / 17 | 41·18 | 8 / 32 | 25·00 |
| August, 2023 | 8 / 21 | 38·10 | 16 / 40 | 40·00 |
| September, 2023 | 11 / 20 | 55·00 | 13 / 34 | 38·24 |
| October, 2023 | 7 / 21 | 33·33 | 15 / 37 | 40·54 |
| November, 2023 | 7 / 18 | 38·89 | 13 / 34 | 38·24 |
| December, 2023 | 10 / 20 | 50·00 | 15 / 44 | 34·09 |
| January, 2024 | 9 / 13 | 69·23 | 18 / 38 | 47·37 |
| February, 2024 | 6 / 16 | 37·50 | 12 / 40 | 30·00 |
| March, 2024 | 9 / 16 | 56·25 | 16 / 32 | 50·00 |
| April, 2024 | 0 / 0 | – – | 0 / 0 | – – |
| May, 2024 | 8 / 13 | 61·54 | 19 / 25 | 76·00 |
| June, 2024 | 9 / 15 | 60·00 | 19 / 27 | 70·37 |
| July, 2024 | 0 / 0 | – – | 0 / 0 | – – |
| August, 2024 | 0 / 1 | 0·00 | 0 / 1 | 0·00 |
| September, 2024 | 4 / 13 | 30·77 | 5 / 19 | 26·32 |
| October, 2024 | 9 / 12 | 75·00 | 15 / 21 | 71·43 |
| November, 2024 | 11 / 16 | 68·75 | 15 / 20 | 75·00 |
| December, 2024 | 10 / 12 | 83·33 | 15 / 21 | 71·43 |

Months without sequencing data are represented as – –. 95% confidence intervals for observed proportions, by month, are illustrated in **Figure S1**.

**Table S7: Effectiveness of doxyPEP against gonorrhea, by *tetM* prevalence (derived from all *N. gonorrhoeae* isolates).**

| Prevalence range of *tetM* or period | Most recent doxyPEP fill | Tests, *n* (%) | | Odds ratio (95% confidence interval) | | Treatment effectiveness (95% confidence interval) |
|---|---|---|---|---|---|---|
| | | Negative | Positive | Unadjusted | Adjusted | Adjusted |
| *tetM* prevalence 20–29·9% | | | | | | |
| | | *N=24,828* | *N=1,300* | | | |
| | Never received | 24,399 (98·3%) | 1,283 (98·7%) | ref. | ref. | ref. |
| | >180d before testing | 208 (0·8%) | 9 (0·7%) | 0·94 (0·61–1·45) | 0·83 (0·53–1·32) | 16·9% (−31·7–47·4) |
| | 91–180d before testing | 84 (0·3%) | 5 (0·4%) | 1·48 (0·87–2·57) | 1·27 (0·72–2·25) | −27·0% (−124·5–28·4) |
| | ≤90d before testing | 137 (0·6%) | 3 (0·2%) | 0·98 (0·64–1·53) | 0·62 (0·38–1·00) | 38·4% (0·3–61·5) |
| *tetM* prevalence 30–39·9% | | | | | | |
| | | *N=25,213* | *N=1,427* | | | |
| | Never received | 24,422 (96·9%) | 1,376 (96·4%) | ref. | ref. | ref. |
| | >180d before testing | 327 (1·3%) | 19 (1·3%) | 1·01 (0·74–1·38) | 0·86 (0·62–1·21) | 13·6% (−21·2–38·1) |
| | 91–180d before testing | 128 (0·5%) | 6 (0·4%) | 1·48 (1·01–2·20) | 1·25 (0·83–1·89) | −24·7% (−88·7–16·8) |
| | ≤90d before testing | 336 (1·3%) | 26 (1·8%) | 1·11 (0·81–1·52) | 0·72 (0·52–1·02) | 27·7% (−1·8–48·3) |
| *tetM* prevalence 40–49·9% | | | | | | |
| | | *N=8,836* | *N=466* | | | |
| | Never received | 8,388 (94·9%) | 427 (91·6%) | ref. | ref. | ref. |
| | >180d before testing | 122 (1·4%) | 9 (1·9%) | 1·09 (0·87–1·35) | 0·90 (0·71–1·14) | 10·1% (−13·7–28·7) |
| | 91–180d before testing | 110 (1·2%) | 16 (3·4%) | 1·48 (1·14–1·94) | 1·22 (0·93–1·62) | −22·5% (−62·3–7·0) |
| | ≤90d before testing | 216 (2·4%) | 14 (3·0%) | 1·26 (1·01–1·56) | 0·85 (0·68–1·07) | 15·1% (−7·0–32·1) |
| *tetM* prevalence ≥50% | | | | | | |
| | | *N=43,209* | *N=2,170* | | | |
| | Never received | 39,571 (91·6%) | 1,923 (88·6%) | ref. | ref. | ref. |
| | >180d before testing | 1,420 (3·3%) | 84 (3·9%) | 1·21 (0·99–1·50) | 0·96 (0·77–1·19) | 4·4% (−18·7–22·8) |
| | 91–180d before testing | 857 (2·0%) | 61 (2·8%) | 1·48 (1·16–1·89) | 1·19 (0·93–1·54) | −19·1% (−53·6–7·0) |
| | ≤90d before testing | 1,361 (3·1%) | 102 (4·7%) | 1·52 (1·26–1·85) | 1·09 (0·89–1·35) | −9·4% (−34·7–10·6) |

We estimated adjusted odds ratios via logistic regression models controlling for: age group; race/ethnicity; transgender identity; commercial or non-commercial insurance source; neighborhood deprivation index; HIV infection status or PrEP receipt; prior receipt of JYNNEOS; prior-year gonorrhea test frequency and history of diagnoses with chlamydia, gonorrhea, or syphilis; and recent (30d) antibiotic receipt. Results of all tests were included, provided individuals did not receive a positive result for the studied pathogen within the preceding 30 days. We define treatment effectiveness as $(1-\mathrm{aOR})\times 100\%$. Analyses allowing effect modification by *tetM* prevalence included 24,542 unique individuals, due to data availability extending only through December, 2024.

**Table S8: Comparison of model fit with differing continuous transformations of effect-modifying covariates.**

| Continuous variable | Transformation | Akaike information criterion | Bayesian information criterion | Area under the curve |
|---|---|---|---|---|
| *tetM* prevalence in GISP *N. gonorrhoeae isolates* | | | | |
| | Linear | 39,408 | 39,667 | 0·723 |
| | Square root | 39,409 | 39,668 | 0·723 |
| | Quadric | 39,411 | 39,689 | 0·723 |
| | Cubic | 39,408 | 39,705 | 0·723 |
| | Log | 39,411 | 39,670 | 0·723 |
| | Logit | 39,409 | 39,668 | 0·723 |
| *tetM* prevalence in all *N. gonorrhoeae isolates* | | | | |
| | Linear | 39,402 | 39,661 | 0·723 |
| | Square root | 39,403 | 39,662 | 0·723 |
| | Quadric | 39,406 | 39,684 | 0·723 |
| | Cubic | 39,402 | 39,699 | 0·723 |
| | Log | 39,404 | 39,662 | 0·723 |
| | Logit | 39,403 | 39,662 | 0·723 |
| Calendar time | | | | |
| | Linear | 39.394 | 39.672 | 0·724 |
| | Square root | 39,397 | 39,675 | 0·723 |
| | Quadric | 39,398 | 39,695 | 0·724 |
| | Cubic | 39,402 | 39,718 | 0·724 |
| | Log | 39,403 | 39,681 | 0·723 |

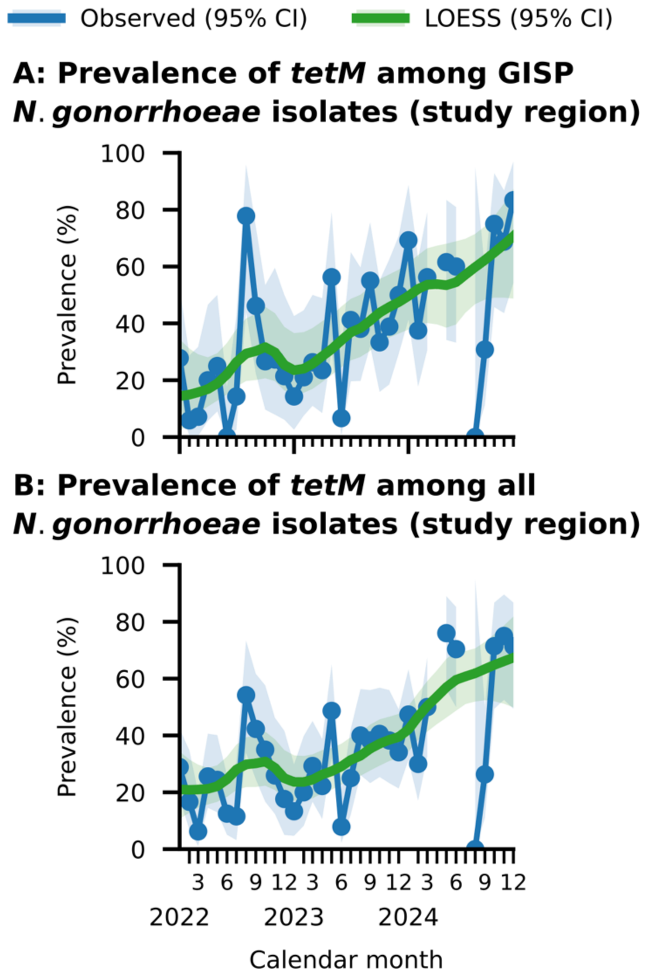


**Figure S1: Prevalence of *tetM* among sequenced gonococcal isolates in the study region, with observed estimates and weighted LOESS smoothing.** We illustrate monthly prevalence of *tetM* and weighted LOESS-smoothed trends in the study region among (**a**) GISP isolates only and (b) all sequenced isolates. We weighted models for LOESS smoothing by the monthly number of isolates collected (**Table S6**). Shaded areas delineate 95% confidence intervals around point estimates (lines).

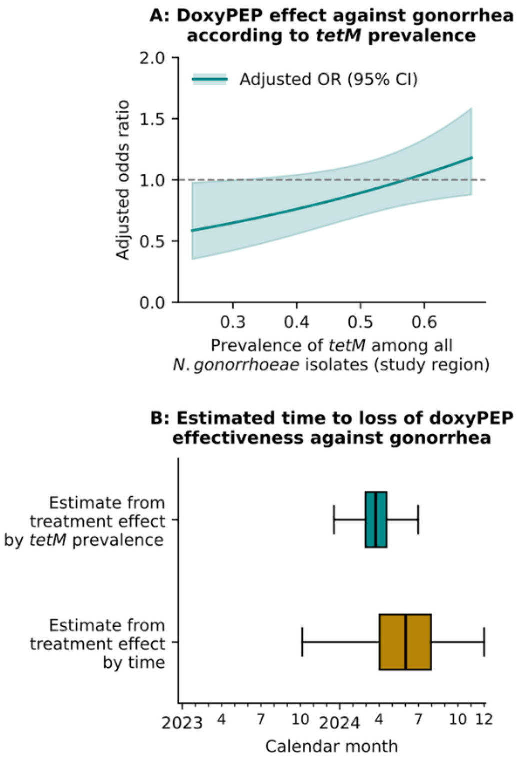


**Figure S2: Variation in doxyPEP effectiveness with *tetM* prevalence among all gonococcal isolates, and time to loss of effect.** We illustrate (**a**) the adjusted odds ratio of a doxyPEP prescription fill in the preceding 90 days among individuals testing positive or negative for *N. gonorrhoeae* infection, defining monthly *tetM* prevalence among all *N. gonorrhoeae* isolates in the study region (GISP and non-GISP isolates) as a linear interaction term based on comparisons of fit for alternative models (**Table S8**). Shaded areas delineate 95% confidence intervals around point estimates. Below, we illustrate (**b**) estimates of the time of loss of doxyPEP effectiveness against gonorrhea, defined as the last date *t* after with aOR(*t*)<1. We project estimates from models defining aOR as a function of *tetM* prevalence (as plotted in panel **a**) and time (as plotted in **Figure 2c**). Shaded areas delineate interquartile ranges around median estimates (lines); outer lines extend to 95% confidence intervals.
